# Polygenic Prediction of Depression Liability Reveals Broad Genetic Associations Across Development, Families, and Populations

**DOI:** 10.64898/2026.11.06.26367433

**Authors:** Tobias Wolfram, Spencer Moore, Jonathan Anomaly, Justin Schleede, Mohammad Ahangari

## Abstract

Polygenic prediction of depression is limited by the depth of depression phenotyping available at biobank scale as well as sample size. Here we develop a depression score combining information from depression and genetically correlated psychiatric traits using phenotype reconstruction, multi-trait genetic integration and Bayesian effect estimation. Compared with the strongest publicly available depression-only score (Psychiatric Genomics Consortium 2025, excluding 23andMe), the liability-*R*^2^ for depression in UK Biobank rose by 49.7% and within-family standardized associations with latent internalizing were up to 54% larger. Across 2,328 childhood and adolescent outcomes in the ABCD study, the association profile of the integrated score was almost identical to that of the depression-only score (r = 0.99), while being uniformly stronger. 1,494 outcomes related to mood, behavior, social functioning, hobbies and sleep were associated with the score — 423 of them within-family. In ABCD, adolescents in the highest score decile were 3.5 times as likely as those in the lowest decile to report suicidal thoughts by age 14 (23% versus 7%) and 1.9 times as likely to report nicotine or cannabis use by age 17 (47% versus 25%). In adults, the score predicted worse general health, chronic pain, and life dissatisfaction. Associations with psychiatric symptoms were substantially stronger in girls than in boys (boys’ coeffiicients 0.68 times girls’), particularly for girl-typical symptoms, and showed no moderation by household resources, hardship or deprivation that remained significant after correction for multiple testing. Prediction was attenuated in line with genetic distance from training data for Hispanic and African American children. Depression prediction can thus be strengthened substantially by integrating the internalizing spectrum, without detectable loss of specificity or within-family predictive ability.

## 1 Introduction

Major depression (MD)^1^ is a common psychiatric disorder and a leading cause of disability world-wide. In 2021, depressive disorders affected approximately 332 million people and accounted for 56 million disability-adjusted life years lost (Rong et al., 2025). Twin-study estimates place the heritability of depression liability at approximately 35–40% (Sullivan et al., 2000) and genome-wide association studies (GWAS) have identified almost 700 associations at more than 600 loci throughout the genome linked to it (Adams et al., 2025).

Polygenic scores (PGSs) are constructed from GWAS summary statistics and can be used to predict individual depression risk, explaining up to 5.8% of MD liability variance in Europeans (Adams et al., 2025). PGSs have been used for the prediction of recurrence and psychiatric comorbidity (Als et al., 2023) and to characterize distinct depression trajectories (Grimes et al., 2024), while phenome-wide studies identified broad adult correlates of depression PGSs and adolescent correlates of cross-disorder genetic liability (Paul et al., 2024; Shen et al., 2020). These applications motivate the development of stronger predictors for research on genetic liability and its broader phenotypic expression.

However, depression is heterogeneous in its clinical presentation. Individuals meeting the same diagnostic criteria can differ substantially in their symptom profiles (Fried and Nesse, 2015). Genetic differences have also been identified between early- and late-onset depression (Shorter et al., 2025), while individual depressive symptoms show different associations with environmental stress (Fried et al., 2015). At biobank scale, detailed depression assessment is often available only for a subset of participants, creating a trade-off between phenotyping depth and sample size (Dahl et al., 2023). Alternative definitions of depression can capture partly distinct genetic architectures, with minimal phenotyping, resulting in genome-wide association signals of low specificity (Cai et al., 2020). Combining the available information can result in higher-quality measures, but complete information is available for only a subset of participants. By combining related measurements, phenotype imputation can improve prediction while preserving specificity to major depressive disorder (MDD) (Dahl et al., 2023).

A related issue is the shared genetic architecture of depression and other psychiatric traits. Depression and anxiety are highly genetically correlated (Thorp et al., 2021). Depression, PTSD and anxiety are associated with an internalizing factor in genomic analyses (Grotzinger et al., 2026). Polygenic scores for depression are associated with the general factor of childhood psychopathology (Waszczuk et al., 2023). Such overlaps provide opportunities for multi-trait analysis of GWAS, which together with modern Bayesian methods and functional genomic annotations can be used to improve polygenic prediction (Turley et al., 2018; Zheng et al., 2024). In this way, depression PGSs can also capture genetic signals that are not specific to MDD (Dahl et al., 2023), which necessitates careful characterisation of shared and trait-specific genetic associations.

Measurement reliability introduces a further consideration. Repeated assessment in female twins demonstrated imperfect reliability of lifetime depression diagnoses (Kendler et al., 1993). Error in the measured phenotype can attenuate PGS associations, motivating comparisons of observed outcomes with estimates that account for measurement reliability (Wolfram et al., 2026). Such estimates depend on the measurement model and should be distinguished from prediction of an observed diagnosis, with both metrics being potentially fit-for-purpose, depending on application.

The interpretation of PGS associations also depends on their persistence within families. Predictions among unrelated individuals can also derive from gene-environment correlations induced by population stratification, indirect genetic effects, and correlations with other genetic factors as a result of non-random mating, including assortative mating. Within-family designs largely isolate direct genetic effects, although some genetic confounding and indirect effects of siblings can remain (Benjamin et al., forthcoming; Chegeni et al., 2026; Huider et al., 2026; Tan et al., 2024; Veller and Coop, 2024; Young et al., 2022). They make it possible to quantify the degree to which polygenic prediction reflects causal effects that vary within families.

Finally, prediction may vary across populations and environmental contexts. Studies have examined sex differences in the genetic architecture of depression and the interplay of depression PGSs and socioeconomic conditions in adolescent outcomes (Campbell et al., 2026; Thomas et al., 2025). Prediction accuracy also varies across the genetic ancestry continuum, with limited transferability of European-derived depression scores to African-ancestry samples (Ding et al., 2023; Kanjira et al., 2025). These findings motivate comparisons not only of individual outcomes, but also of whether the broader pattern of associations differs across groups.

Here, we develop an integrated depression PGS using phenotype information from multiple large-scale biobanks together with external GWAS summary statistics. The score combines information from depression and genetically correlated psychiatric traits using phenotype reconstruction, multi-trait genetic integration and Bayesian effect estimation, while retaining depression as the focal prediction target (Methods). On a common scale, the integrated summary statistics carry about twice the genome-wide association signal of the PGC depression GWAS (Results).

We evaluate prediction in UK Biobank and the Adolescent Brain Cognitive Development (ABCD) Study and contrast it with a recent best-in-class literature PGS developed by the PGC 2025 European-ancestry meta-analysis excluding 23andMe, the largest publicly available depression summary data (Adams et al., 2025). We first compare our summary data with that of the PGC, then examine prediction of depression and the role of outcome reliability, followed by associations with latent internalizing psychopathology, extending these analyses to physical and mental health, along with behavioral, cognitive, and socioeconomic outcomes. Finally, we examine sex differences, interactions with measured family environments and cross-ancestry portability. Together, these analyses investigate how well the score predicts MD and the overall depression-adjacent phenome as well as the context-dependency of these predictions.

## 2 Results

### 2.1 Integration strengthens the genome-wide depression signal

We compared the integrated discovery summary statistics with those of the published PGC GWAS (without 23andMe) on HapMap sites. The integrated GWAS carried 2.10 times (95% CI 2.04– 2.17) the genome-wide association signal of the PGC GWAS, measured as the ratio of LD-score regression slopes, which scale with sample size times SNP heritability (Bulik-Sullivan et al., 2015). The increase was uniform across the LD-score distribution (2.07–2.16 across quintiles), and the two sets of statistics were almost perfectly genetically correlated (*r_g_*= 0.98).

### 2.2 The MD PGS predicts depression, internalizing and adult life and health outcomes in population and family analyses

We defined an MD reference phenotype in our UK Biobank validation sample using the published LifetimeMDD definition (Cai et al., 2020; Dahl et al., 2023) and compared our integrated predictor’s performance with the PGC benchmark (Methods). Classifications often changed on reassessment. Of 11,567 UK Biobank participants initially classified as cases and reassessed after a median of 6.2 years, only 47.7% remained cases, whereas 38.9% moved to a symptom-positive unclassified category and 13.5% were classified as controls. Treating these changes as measurement error, we report approximate liability-scale estimates before and after a model-based repeat-stability correction (Methods, Supplementary Table S3C).

The approximate population liability-*R*^2^ was 7.0% for the integrated PGS and 4.6% for the PGC score before correction, and 9.1% and 6.1% after correction, a relative gain of 49.7% (95% CI 27.6–78.0%, Table 1, Supplementary Table S3A and B). To assess within-family prediction, we fitted SNIPar models with parental scores imputed from genotyped siblings (Guan et al., 2025; Young et al., 2022) in 7,054 participants from 6,085 sibships^2^. The direct/population coeffiicient ratio was 1.03 (95% CI 0.83–1.23) for the integrated score and 1.02 for the PGC score (Table 1), so there was no detectable within-family attenuation, although reductions of up to about 17% cannot be excluded. Applying these ratios to the population estimates gave model-implied within-family liability-*R*^2^ of 9.6% and 6.3%, respectively (Figure 1A). Matched observed-scale results confirmed this picture, and the integrated PGS also explained more observed variance in anxiety symptoms, neuroticism and internalizing than the PGC score, with relative gains (83–94%) larger than for depression itself (51%), although with a wide interval (Supplementary Table S4). In ABCD, which contributed no discovery data, the relative gain for depression diagnosed with the computerized Kiddie Schedule for Affective Disorders and Schizophrenia (K-SADS) in childhood and adolescence was similar (liability-*R*^2^ 4.9% versus 3.5%, a 39% gain, 95% CI 17–66%), although this construct differs from adult lifetime depression (Supplementary Note 2).

**Figure 1:**
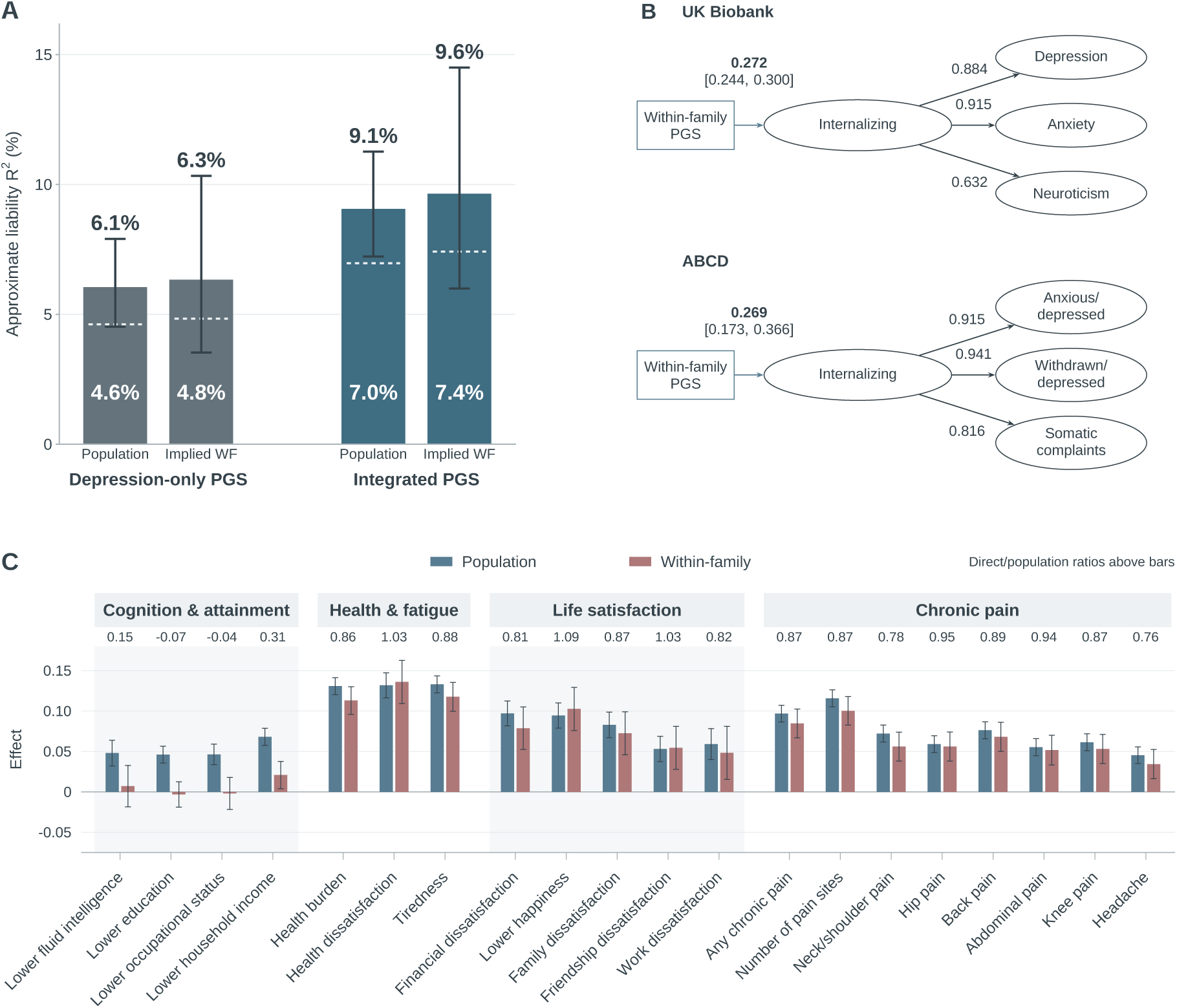
Depression, internalizing, and adult family associations. **A,** Bars and labels above the whiskers show approximate liability-*R*^2^ while correcting for measurement error, with 95% CIs. White labels/dashed marks give the estimates before correction. Within-family liability-*R*^2^ values are model-implied (population value *×* squared SNIPar direct/population ratio), and their intervals include uncertainty in the ratio from 4,000 whole-family bootstrap draws. **B,** Standardised within-family associations with latent internalizing (sibling difference estimate) in UK Biobank and ABCD. **C,** Standardised population and direct (parental-score-adjusted) SNIPar associations of the integrated score with adult outcomes in the UK Biobank sibling validation sample. Numbers above the bars give direct/population ratios. Intervals are 95% CIs.

**Table 1:** UK Biobank depression: population and within-family prediction by the integrated and PGC scores.

|  | Integrated | PGC |
|---|---|---|
| Population liability-*R*<sup>2</sup>, uncorrected (%) | 7.0 (5.5–8.7) | 4.6 (3.4–6.1) |
| Population liability-*R*<sup>2</sup>, repeat-corrected (%) | 9.1 (7.2–11.3) | 6.1 (4.5–7.9) |
| Direct/population ratio | 1.03 (0.83–1.23) | 1.02 (0.78–1.26) |
| Model-implied within-family liability-*R*<sup>2</sup>, repeat-corrected (%) | 9.6 (6.0–14.5) | 6.3 (3.5–10.3) |

*Note.* 7,054 participants (1,679 cases) from 6,085 sibships of the sibling validation sample. Liability-*R*<sup>2</sup> assumes a population prevalence of 15%, and the repeat correction uses the polychoric reliability (Supplementary Table S3D). Direct/population ratio is derived from linear mixed models of standardized case status on the parental-compatible SNIPar score, without (population) and with (direct) adjustment for parental scores imputed from siblings. Intervals are 95% intervals from 4,000 paired whole-family bootstrap draws.

To study the broader mood–anxiety spectrum, we modelled latent internalizing, a hierarchical factor capturing variance shared across depression, anxiety and neuroticism items in UK Biobank (28 PHQ-9, GAD-7 and EPQ-R items) and across anxious/depressed, withdrawn/depressed and somatic-complaint items in ABCD (30 Youth Self-Report items), and estimated within-family PGS associations from sibling differences in the score (Methods). The integrated PGS coeffiicients were 27.8% (95% CI 17.4–39.2%) higher than those for PGC in UK Biobank (*β* = 0.272 versus 0.213) and 54.2% (8.5–119.2%) higher in ABCD (*β* = 0.269 versus 0.175, Figure 1B and Supplementary Table S6), and the gains were essentially unchanged under alternative factor models. Within-family attenuation was similar for the two scores in UK Biobank, whereas in ABCD the integrated score retained a larger share of its population association with internalizing (by 37.3 percentage points, 95% CI 4.2–70.3, Supplementary Table S7).

We also observed broad associations with poorer general health, tiredness, life dissatisfaction and chronic pain in the UK Biobank sibling validation sample (10,387–36,257 participants depending on outcome, Figure 1C). Most of these associations remained within families, with direct/population ratios of 0.86 (95% CI 0.76–0.97) for general health burden and 0.87 (0.72–1.02) for chronic pain. The socioeconomic and cognitive associations, by contrast, were substantially attenuated, with ratios of 0.07 for lower education, 0.04 for lower occupational status, 0.15 for lower fluid intelligence and 0.31 for lower household income. Patterns were similar for the PGC score (Supplementary Note 4).

Associations with 23 diagnoses across psychiatric, cardiometabolic, immune and inflammatory, cancer and other disease categories (Table 2) were more limited: direct within-family associations after Benjamini–Hochberg correction (Benjamini and Hochberg, 1995) remained significant for just bipolar disorder, coronary artery disease, hypertension, type 2 diabetes, venous thromboembolism and asthma, with direct/population ratios significantly below 1 for hypertension (0.64) and type 2 diabetes (0.60). Results were directionally similar for the PGC score (Supplementary Data).

**Table 2:**
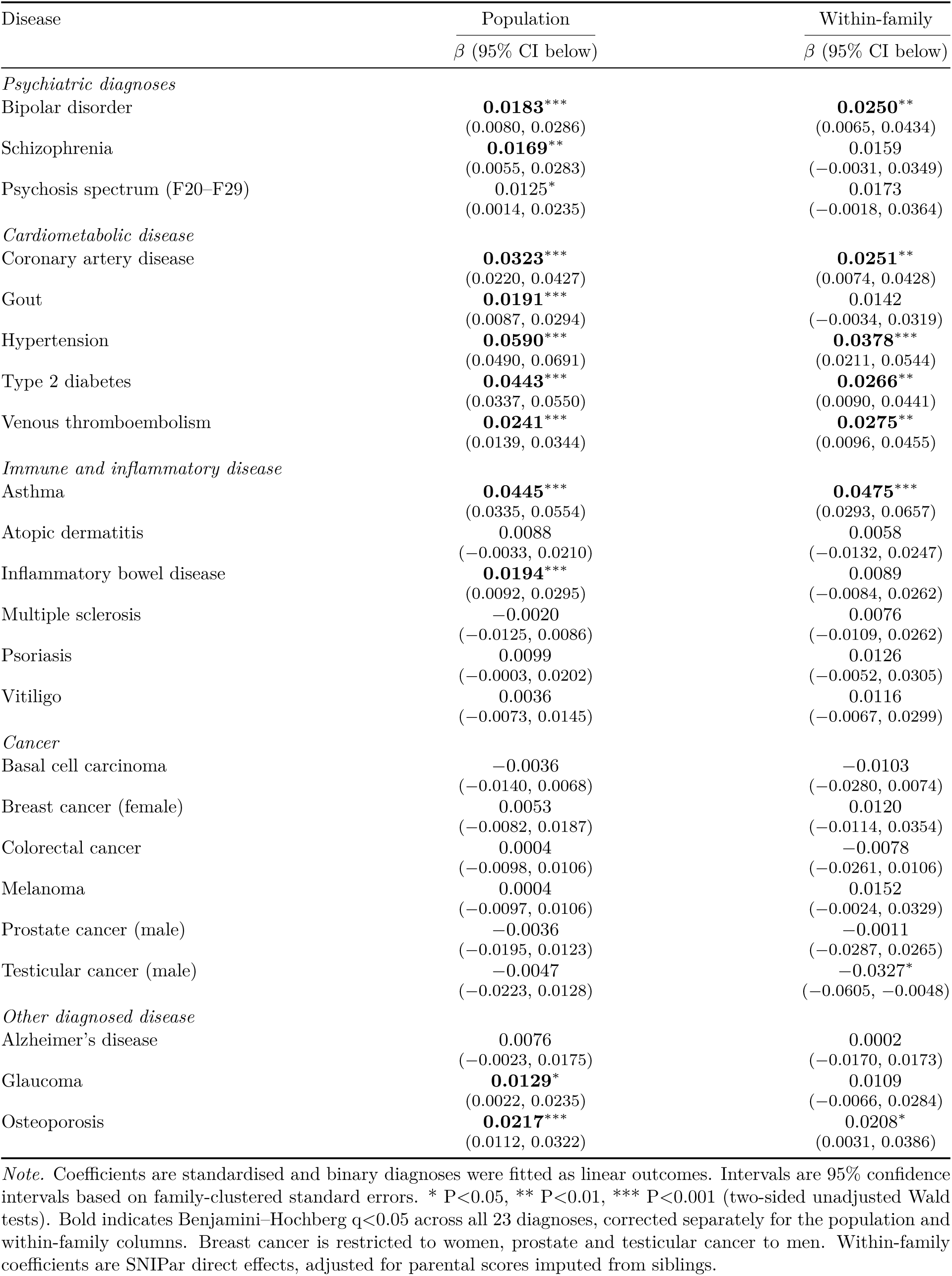
Population and within-family associations of the integrated depression score with disease outcomes in UK Biobank.

### 2.3 Depression liability reveals broad associations across the childhood phenome

The rich data available in ABCD enabled us to study early-life correlates of the integrated PGS across 2,328 outcomes from 28 instruments concerning social functioning, behavior and self-regulation, mood, stress and psychiatric symptoms, and activity, recreation and sleep. Of these, 1,494 were associated with the PGS after Benjamini–Hochberg correction across the 2,328 out-comes (q<0.05), including 423 with nominally significant and directionally similar within-family associations (P<0.05, Figure 2A). The profile extended to emotional dysregulation, bullying and peer diffiiculties, attention and behavioral problems, and sleep disturbance. Supplementary Data gives all outcomes and estimates.

**Figure 2:**
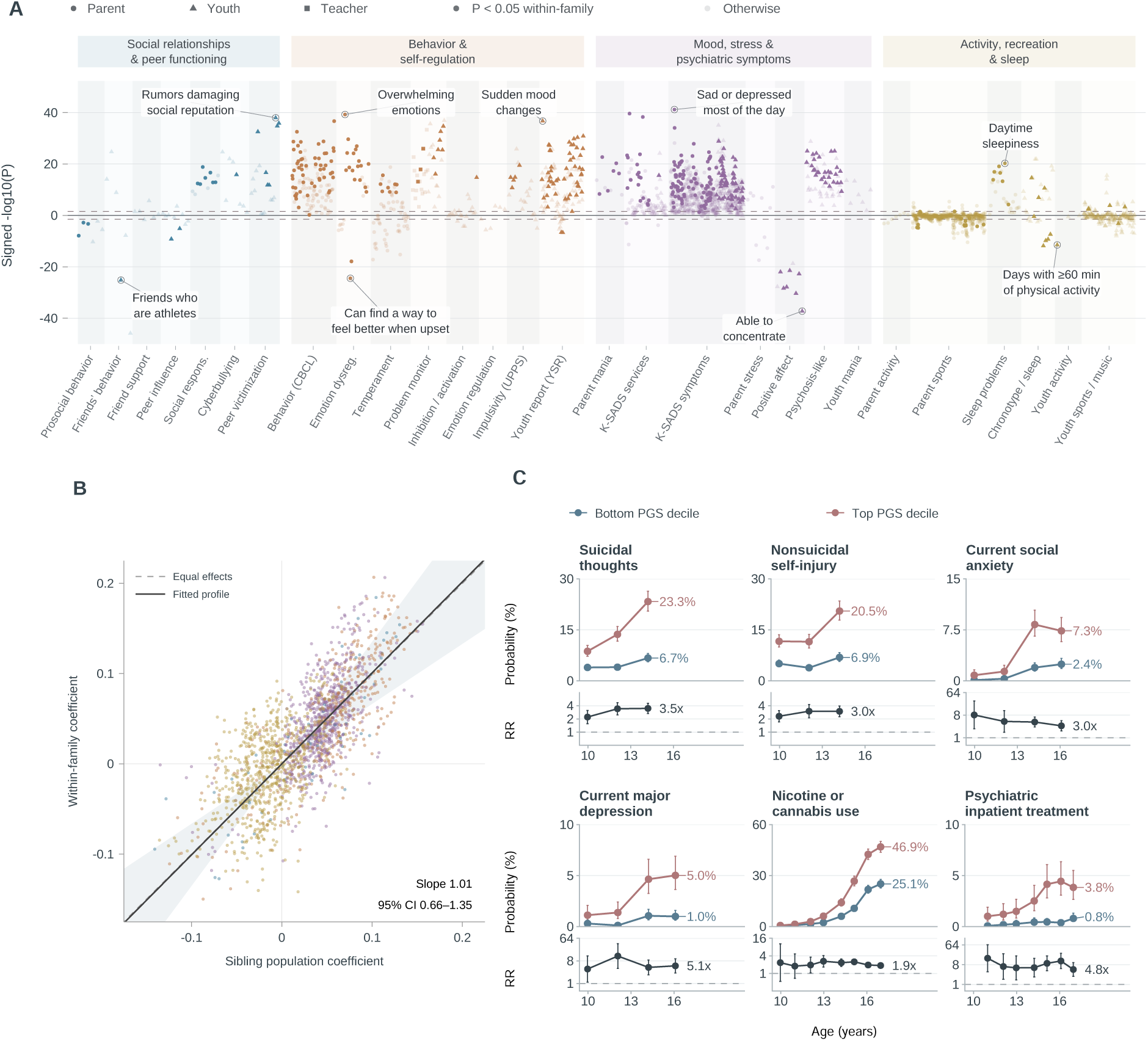
Childhood phenome and developmental trajectories. **A,** Signed population association strength across 2,328 outcomes. Colors identify domains and symbols distinguish reporters. More opaque points have same-direction nominally significant within-family associations (P<0.05). Dashed lines show the BH FDR 0.05 cutoff across the displayed outcomes. **B,** Matched within-family and population coeffiicients, with the covariance-corrected profile slope and 95% interval. The dashed line denotes equal effects. **C,** Model-standardized probabilities at the top and bottom PGS deciles across assessments, with corresponding risk ratios and pointwise 95% intervals. Suicidal thoughts and self-injury refer to reported past/lifetime history and nicotine or cannabis use to lifetime use, inpatient treatment to the period since the preceding assessment. Supplementary Note 5 describes the models. Supplementary Data provide the PheWAS associations.

For the 1,854 outcomes measurable in the subset of siblings and dizygotic twins within ABCD, the errors-in-variables slope of within-family on population coeffiicients across outcomes was 1.01 (95% CI 0.66–1.35, Figure 2B). On average, the profile was therefore carried by direct effects, although the interval allows substantial attenuation or amplification.

Against the PGC score, the mean incremental observed-scale *R*^2^ was 24.7% higher (95% CI 10.8–41.4%, Supplementary Table S4), though the within-family *R*^2^ gain was uncertain (12.3%, Supplementary Data). Nevertheless, the two scores produced closely aligned population association profiles across all 2,328 outcomes in the childhood phenome (*r* = 0.988, Supplementary Table S10). The PGC score retained somewhat less of its population association profile within families (slope 0.90 versus 1.01), but the paired difference was not significant (Supplementary Table S7).

For six illustrative outcomes, we estimated PGS associations across assessments at approximately ages 10–17 (Figure 2C). By the last assessment (age 14 for suicidal thoughts and self-injury, 16 for depression and social anxiety, 17 for the others), children in the highest PGS decile were more likely than those in the lowest to report suicidal thoughts (23.3% versus 6.7%, RR 3.48), nonsuicidal self-injury (20.5% versus 6.9%, RR 2.98), current depression (5.0% versus 1.0%, RR 5.07), current social anxiety (7.3% versus 2.4%, RR 3.01), nicotine or cannabis use (46.9% versus 25.1%, RR 1.87) and psychiatric inpatient treatment since the preceding assessment (3.8% versus 0.8%, RR 4.82). Larger absolute probabilities did not always imply larger relative associations: risk ratios for social anxiety and inpatient treatment were larger, though less precise, at earlier assessments.

### 2.4 Sex differences in symptom associations and household moderation

We next asked whether the strength and expression of depression-related genetic liability varied by sex and household context in ABCD.

Among the 726 outcomes in the selected psychiatric symptom batteries, 107 showed sex differences after Benjamini–Hochberg correction across these 726 tests. Associations were stronger in girls for 101 outcomes and in boys for six (Figure 3A). The contrasts more positive in girls included fatigue, guilt, low self-esteem, indecision, panic and uncontrollable worry, and self-injury. Those more positive in boys primarily concerned conduct problems (physical fights, bullying, breaking and entering, vandalism and cruelty to animals). Supplementary Data provides all sex-specific coeffiicients and contrasts.

**Figure 3:**
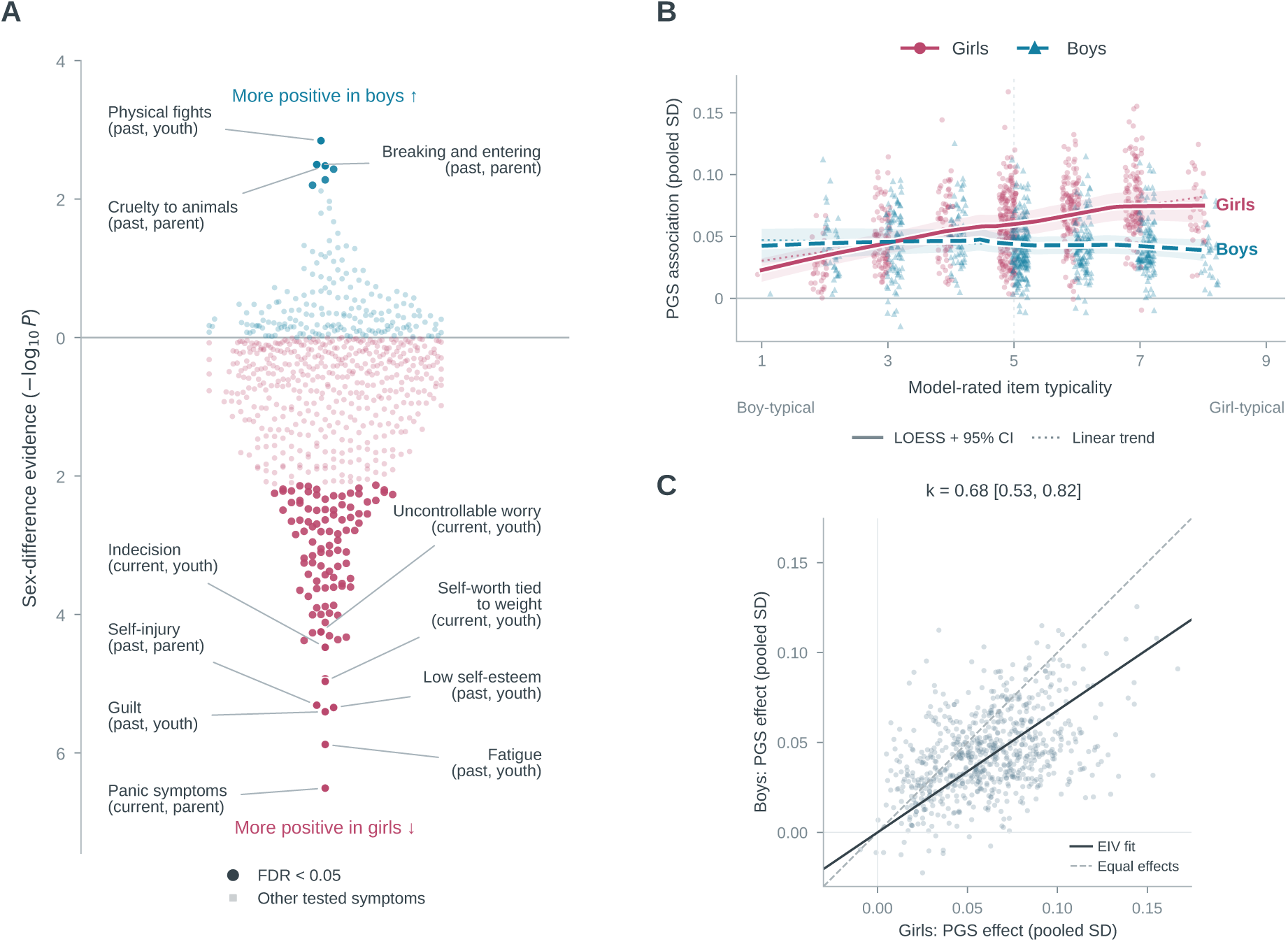
Sex-specific PGS associations across psychiatric symptoms in ABCD. **A,** Sex contrasts in the selected psychiatric symptom batteries, with signed vertical position given by sign(*β*_male_ *− β*_female_) *×* [*−* log_10_(*P*)] for a two-sided sex-difference test. Blue denotes a more positive male coeffiicient and rose a more positive female coeffiicient. This contrast does not necessarily indicate a larger absolute association. Darker, larger points pass Benjamini– Hochberg FDR <0.05 across the 726 displayed outcomes. **B,** Relationship between cultural gender typicality of symptom content and sex-specific PGS associations. A language model rated items from boy-typical to girl-typical using their descriptions, reporter and response options, without access to participant data or association results. Rose circles and the solid curve denote girls, blue triangles and the dashed curve denote boys. Curves are local linear LOESS fits with pointwise 95% confidence bands. Dotted lines show equal-item-weighted linear trends. Coeffiicients are standardised. The panel includes 716 rated items. Ten outcomes without usable ratings remain in A and C. **C,** Girls’ and boys’ coeffiicients for the same outcomes as A. The dashed line indicates equality and the solid line the corrected-moment errors-in-variables slope, *k*, with its 95% interval. See Supplementary Note 6 for selection, ratings, scaling and uncertainty.

Given these differences, we examined in an exploratory analysis whether sex-specific PGS associations varied with the cultural gender typicality of symptom content. A language model rated each item from strongly boy-typical to strongly girl-typical using only its wording or description, reporter and response options. Among the 716 items with usable ratings, associations in girls increased with girl-typicality, whereas boys showed little linear trend (Figure 3B). Per one-point increase in typicality, the PGS coeffiicient increased by 0.0074 pooled outcome SD in girls and changed little in boys (between-sex difference 0.0082, 95% CI 0.0050–0.0114). Because these coeffiicients combine differences in association and symptom dispersion, within-sex outcome standardization shrank the difference (Supplementary Data).

Across the symptom profile, boys’ coeffiicients were 0.68 times those in girls (95% CI 0.53–0.82), corresponding to approximately 32% smaller associations on average (Figure 3C). After within-sex outcome standardization, the difference shrank to 21%. Results were similar for the whole phenome analysis and for the PGC benchmark score (Supplementary Note 6). In UK Biobank adults, the score’s liability-*R*^2^ for the reference depression definition was likewise higher in women than in men (repeat-corrected 9.9% versus 8.2%), but the difference was not statistically significant (1.7 percentage points, 95% CI *−*2.7 to 5.8).

In contrast, household resources, hardship and residential deprivation showed no clear moderation of latent internalizing or either the symptom or full PheWAS profile (Table 3). For the symptom profile, point estimates suggested somewhat stronger associations under disadvantage, but none remained significant after Holm correction (Holm, 1979) across the three exposures (smallest adjusted *P* = 0.08). Latent internalizing interactions were close to zero (at most 0.016 latent SD per exposure SD) and did not differ detectably between the two scores (Supplementary Note 6).

**Table 3:** Sex and household moderation of latent internalizing, psychiatric symptoms and the full PheWAS profile in ABCD.

| Moderator | Internalizing<br>Interaction (latent SD) | Symptom profile<br>Profile moderation | Full PheWAS profile<br>Profile moderation |
|---|---|---|---|
| Sex (boys minus girls) | −0.099<br>[−0.151, −0.047] | −0.322<br>[−0.473, −0.177] | −0.292<br>[−0.430, −0.158] |
| Resources | −0.007<br>[−0.039, 0.025] | −0.145<br>[−0.306, 0.012] | −0.102<br>[−0.248, 0.045] |
| Hardship | 0.007<br>[−0.021, 0.035] | 0.163<br>[0.017, 0.307] | 0.108<br>[−0.018, 0.229] |
| Residential deprivation | 0.016<br>[−0.026, 0.059] | 0.108<br>[−0.080, 0.295] | 0.107<br>[−0.063, 0.274] |

Estimates with 95% CIs below. Household exposures are standardized. Profile estimates are boys-to-girls multipliers minus one (sex) or slopes of interaction coefficients on the covariate-averaged main PGS coefficients (household exposures). No household profile slope was significant after Holm correction across the three exposures. The symptom profile covers 726 outcomes (Figure 3). The full profile includes all eligible PheWAS outcomes.

### 2.5 Core associations and phenotype profiles across population groups

Lastly, we compared PGS performance across population groups, first for depression and latent internalizing and then across the childhood phenome.

In UK Biobank, the integrated PGS association with depression was lower than the European reference by 61.9% in the African-ancestry (AFR) group, 71.2% in East Asians (EAS) and 15.7% in South Asians (SAS, Figure 4A). Intervals were wide because of small samples and included published expected effect reductions of 57.6%, 30.3% and 19.6%, respectively (Privé et al., 2022) (Supplementary Note 7 and Table S12A).

**Figure 4:**
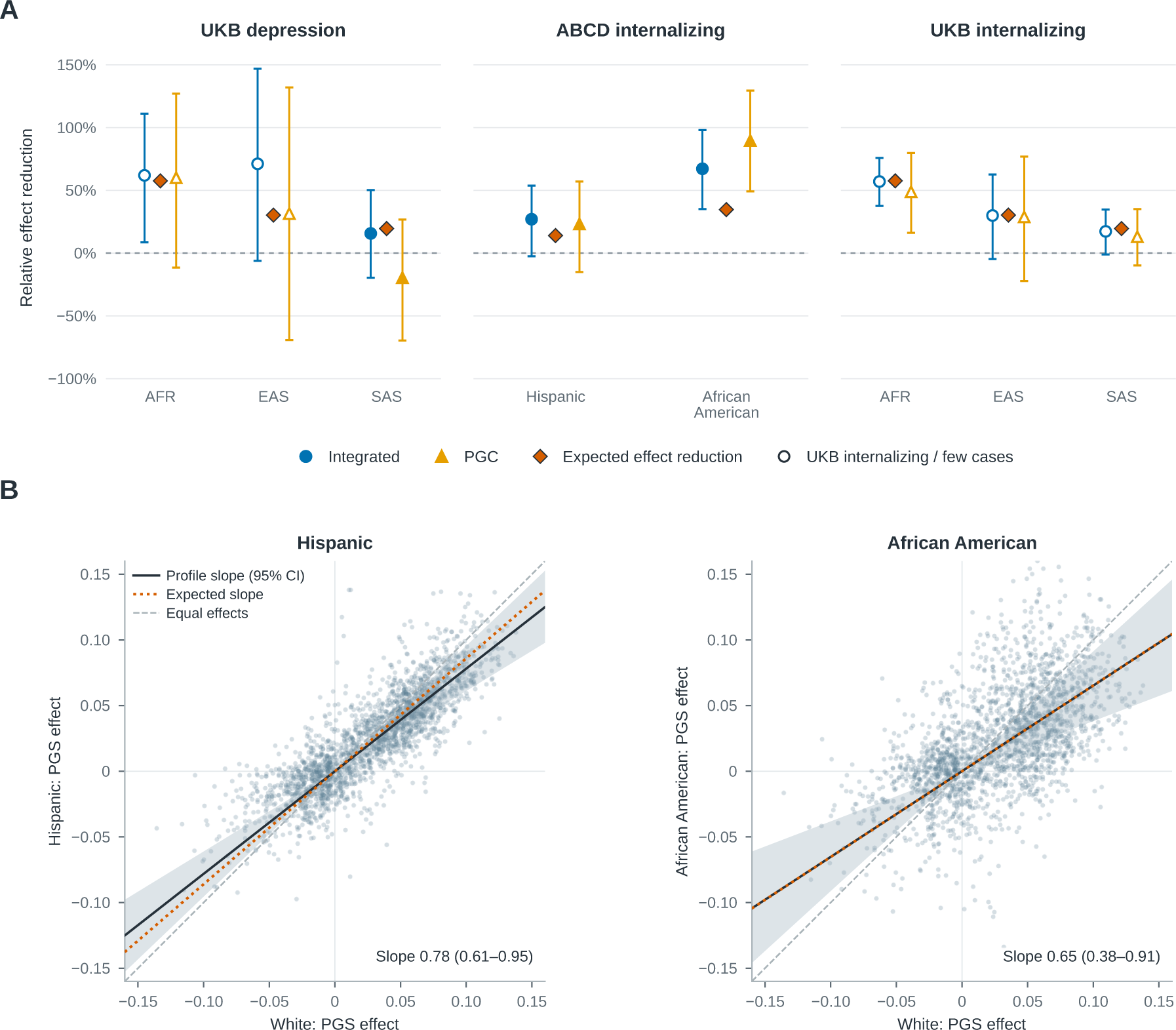
Core and phenotype-wide portability. **A,** Relative reduction, 1 *− β_g_*/*β*_ref_, for UK Biobank depression and ABCD and UK Biobank internalizing. Positive values indicate smaller associations and negative values larger associations than the reference group (EUR in UK Biobank, White children in ABCD). Circles denote the integrated PGS and triangles the PGC benchmark. Depression uses log-odds coeffiicients. Internalizing coeffiicients are standardised. Bars show 95% intervals. Open markers identify the UK Biobank internalizing analysis and AFR/EAS diagnosis cells with few cases. **B,** ABCD phenotype-wide associations. Solid lines show errors-in-variables slopes. Shaded bands describe pointwise 95% CIs. Dashed lines denote equal effects. The view is cropped. The complete plotting range is shown in Supplementary Figure S1. Diamonds in A show expected effect reductions and dotted lines in B expected profile slopes, derived from published relative *R*^2^ estimates for UK Biobank (Privé et al., 2022) and ABCD (Moore et al., 2025; Wolfram et al., 2026). See Supplementary Note 7 for sample, model, measurement-comparability and benchmark limitations.

In ABCD, the integrated-score association with latent internalizing was reduced by 27.0% in the Hispanic group and 67.2% (95% CI 35.1–98.1%) in the African American group, compared with published expected reductions of 14.0% and 34.7%, respectively (Moore et al., 2025; Wolfram et al., 2026) (Supplementary Table S12A).

Across the ABCD phenome, the slope relative to White children was 0.78 (95% CI 0.61–0.95) in Hispanic children and 0.65 (0.38–0.91) in African American children (Figure 4B), indicating approximately 22% and 35% attenuation in the overall coeffiicient profiles, compared with the same expected reductions of 14.0% and 34.7%. Supplementary Figure S1 shows the full coeffiicient range. The PGC score showed a similar pattern, with slopes of 0.79 and 0.58 (Supplementary Table S12B).

## 3 Discussion

We developed a depression polygenic score that outperforms the strongest publicly available depression score, based on the European-ancestry PGC 2025 summary statistics excluding 23andMe (Adams et al., 2025). By integrating observed and reconstructed depression phenotypes from multiple biobanks with external broader psychopathology spectrum GWAS, we obtained summary statistics carrying about twice the genome-wide association signal of the PGC GWAS. In UK Biobank, the approximate liability *R*^2^ for depression rose from 4.6% to 7.0% (a relative increase of 51%) and, after correction for repeat-measurement error, from 6.1% to 9.1% (49.7%). Within families, standardized coeffiicients with latent internalizing were 27.8% larger than those of the benchmark score in UK Biobank and 54.2% stronger in ABCD, with gains extending to neuroticism and anxiety.

A common concern with integrating broader phenotypes and genetically correlated traits is that power is gained at the cost of specificity, so that the score increasingly reflects anxiety, neuroticism or similar traits rather than depression (Cai et al., 2020; Dahl et al., 2023). A score that had drifted toward the internalizing spectrum would gain least on strict depression and most on internalizing-type outcomes. The integrated score did gain on depression itself, in population and within-family analyses alike. In the population analyses, however, the gains for anxiety, neuroticism and the internalizing composite were larger than for depression (83–94% versus 51% in incremental *R*^2^, corresponding to roughly 35–39% versus 23% on the coeffiicient scale, Supplementary Table S4). Within families, by contrast, the gains were of the same order for all four outcomes (20–33% for anxiety, internalizing and neuroticism, Supplementary Table S6, and about 24% for depression, with direct coeffiicients of 7.74 versus 6.25 percentage points per SD, Supplementary Note 2). Neuroticism showed the largest within-family gain (33%), so some reweighting toward neuroticism cannot be excluded, although the depression estimate is too imprecise to distinguish this from a uniform gain. Across the childhood phenome the gain was proportional: the association profiles of the two scores were closely aligned across all 2,328 ABCD outcomes (*r* = 0.988), and the integrated score’s coeffiicients were on average 11% larger and its mean incremental *R*^2^ 24.7% higher in the 1,854-outcome common inventory (Supplementary Tables S4 and S10). The integrated score thus strengthened the associations the depression-only score already had rather than adding new ones. Together, these results limit the concern about specificity but do not rule it out.

Beyond the comparison with PGC, the score predicts not only depression but a broad liability to mood and anxiety problems, both in the population as well as within-family. Symptom dimensions of this kind capture liability that a lifetime diagnosis, which compresses it into two imperfectly reliable categories (Kendler et al., 1993), does not.

The score also predicted a wide range of adult outcomes. Associations with poor health, tiredness, chronic pain and life dissatisfaction were retained within families. By contrast, associations with fluid intelligence, education and occupational status were reduced to (almost) zero within families, and the association with household income became barely significant. For socioeconomic outcomes, population stratification, assortative mating, and/or indirect genetic effects are therefore likely to account for much of the population association. Because ascertainment in UK Biobank also shifts genetic correlations between depression and cognition (Nguyen et al., 2022; Tyrrell et al., 2021), the cognitive associations in particular should be interpreted with caution. Disease associations were smaller on the standardized scale, and their within-family point estimates were often lower. The absence of detectable within-family attenuation for depression (direct/population ratio 1.03, 95% CI 0.83–1.23) agrees with twin and parent–offspring studies of depression polygenic scores (Huider et al., 2026; Trindade Pons et al., 2025), although our interval does not exclude attenuation of up to about 17%. Genome-wide, family-based GWAS estimate a direct–population genetic correlation of only 0.68 for depression, indicating substantial confounding of population effects, although population effects were on average smaller rather than larger than direct effects (Tan et al., 2024), in contrast to the within-sibship shrinkage reported for strongly associated variants for depressive symptoms (Howe et al., 2022).

In a phenome-wide scan of 2,328 outcomes in ABCD, 1,494 were associated with the score after correction for multiple testing. Higher scores were associated with greater peer victimization, including reputation-damaging rumors, greater diffiiculty regulating emotions, more sleep problems and less physical activity. These associations retained their strength within families on average. Children in the top score decile had substantially elevated probabilities of suicidal thoughts and self-injury and were about five times as likely as children in the bottom decile to have received psychiatric inpatient treatment since the preceding assessment. These results extend earlier work on adolescent depression trajectories and on the childhood correlates of internalizing liability (Grimes et al., 2024; Paul et al., 2024).

Household resources, material hardship and residential deprivation did not detectably moderate the score’s association with latent internalizing, in line with the minimal interaction between depression PGS and household income reported in ABCD (Campbell et al., 2026). Across the psychiatric symptom profile, point estimates suggested somewhat stronger associations under disadvantage, but none remained significant after correction across the three exposures. Sex, by contrast, mattered substantially. Across 726 psychiatric symptom outcomes, a corrected-moment profile slope placed boys’ coeffiicients at around 32% less than girls’, a pattern also evident for the PGC score on the same psychiatric outcomes and across the broader phenome. Symptoms with significant sex differences almost always had more positive coeffiicients in girls, including panic symptoms, fatigue, guilt, low self-esteem and self-injury.

The sexes may also differ in which symptoms express the liability. In girls, associations increased with the rated girl-typicality of symptoms, whereas in boys they were largely independent of it. With sex-specific standardization, however, the difference between the two trends attenuated, so the gradient depends substantially on sex differences in symptom variance, which for binary and near-binary items largely reflects sex differences in symptom prevalence. The overall pattern aligns with the adolescent onset of female excess in depression (Salk et al., 2017), though, as male and female depression share most of their genetic architecture (Thomas et al., 2025), the weaker association in boys does not require different causal variants and can be the result of differences in symptom expression, reporting or prevalence, including the hormonal changes of puberty (Angold et al., 1999).

Cross-ancestry effect reductions were in line with expectations for a score trained on European-ancestry data (Ding et al., 2023; Kanjira et al., 2025; Wang et al., 2020). Although intervals were wide, almost every estimate was compatible with published portability expectations for other traits (Moore et al., 2025; Privé et al., 2022). Closing the portability gap will require more diverse discovery data.

Taken together, the integrated score predicts depression more strongly and captures more of the internalizing spectrum, including neuroticism, while in its general phenome-wide association profile remaining closely aligned with that of the depression-only benchmark score and showing minimal within-family effect reduction. For depression, broad phenotypic integration across the internalizing spectrum could therefore roughly double the association signal of a depression GWAS without evidence of the loss of specificity that has been a concern for minimally phenotyped GWAS (Cai et al., 2020).

Several limitations apply. First, the PGC benchmark excluded 23andMe, which contributed about 113,000 European-ancestry cases and 1.77 million controls, an effective sample size of about 425,000 against about 1.15 million for the European meta-analysis without it (Adams et al., 2025). A score including these data would be a stronger benchmark, but the same data could be added to our framework, and phenotype reconstruction would plausibly benefit from 23andMe’s survey data as well. Second, the integrated score’s effects were estimated with SBayesRC, which uses functional annotations, whereas the benchmarks used SBayesR, as in the PGC study. Part of the score-level gain may therefore reflect the estimation method rather than phenotype reconstruction and multi-trait integration. The comparison of the summary statistics themselves, which precedes effect estimation, is unaffected. Third, the measurement-error correction treats disagreement between two questionnaires as error, although it may also reflect new episodes or changed recall (Kendler et al., 1993; Moffiitt et al., 2010). Under a liability framing, however, a single assessment is still an imperfect indicator of a stable liability. Fourth, participation bias can distort genetic associations in UK Biobank (Schoeler et al., 2023), and the ABCD family sample and the non-European samples were small.

## 4 Methods

### 4.1 Participants and study design

For validation, we used UK Biobank and ABCD data (ABCD Study, 2026; Sudlow et al., 2015). We confirmed no overlap between local discovery GWAS and UK Biobank validation samples. External GWAS inputs to the integrated score excluded UK Biobank, while ABCD contributed no discovery data.

#### UK Biobank sample

UK Biobank validation used genotyped sibships, with family relationships identified using KING (Manichaikul et al., 2010). Family validation analyses used participants in the United Kingdom genetic-ancestry group (Privé et al., 2022), excluding heterozygosity/missingness outliers and sex-chromosome aneuploidies.

In line with the reference phenotype construction (Supplementary Note 1), participants in the analysis in Figure 1A additionally required British self-reported ethnicity, the UK Biobank genetic ethnic grouping labelled “Caucasian,” and agreement between reported and genetic sex. The sample comprised 7,054 participants. Repeat-assessment corrections used respondents with interpretable classifications at both assessments and were not restricted to the family cohort. The latent internalizing analysis required at least two eligible siblings per family, each with at least one observed symptom item, leading to 36,296 participants from 17,656 families. Adult SNIPar analyses used 10,387–36,316 participants from 7,776–17,668 sibships of the sibling validation sample, depending on outcome. Supplementary Note 2 specifies the depression case/control rules, and cross-population samples are described under Population portability.

#### ABCD sample

We used phenotypes from Release 7, with consortium-processed genotypes and genetic/demographic metadata. Primary family analyses used parent-reported White siblings and dizygotic twins. Monozygotic twins and twins with missing or conflicting zygosity were excluded. The latent-family sample contained 1,172 participants in 593 families. Broader population and portability analyses used separate eligible samples, with the broader White population including singletons, siblings and twins (Supplementary Notes 1 and 5–7). Score comparisons matched participants and covariates within each analysis. Reference score units remained fixed after outcome restriction and resampling (Supplementary Note 1).

### 4.2 Polygenic score construction and benchmarks

#### Scope of disclosure

This manuscript describes the evaluation of an integrated depression predictor and comparisons with literature-based GWAS benchmarks. Detailed predictor construction methods and input specifications are withheld in this version and will be disclosed in the published paper. The validation methods and benchmark specifications are described here and in the Supplement.

The integrated depression PGS combined GWAS of observed and reconstructed depression phenotypes from multiple biobanks with external GWAS summary statistics using multi-trait analysis of GWAS (MTAG) (Turley et al., 2018). Posterior SNP effects were subsequently estimated using SBayesRC in GCTB 2.5.2 (Zheng et al., 2024). To compare the integrated GWAS with the published PGC GWAS without 23andMe, we used ratios of LD-score regression slopes on identical variants (Bulik-Sullivan et al., 2015). These slopes are proportional to sample size times SNP heritability and are estimated from the association statistics alone (Supplementary Note 1).

#### PGC benchmarks

Depression-only comparative analyses used the European PGC MDD2025 release without 23andMe (Adams et al., 2025) and, in line with the PGC study, SBayesR (Lloyd-Jones et al., 2019) for score construction. ABCD used the published UK Biobank-inclusive GWAS. All UK Biobank analyses used PGC excluding UK Biobank combined with a local depression GWAS excluding evaluation participants and their relatives. The local phenotype was designed to approximate PGC depression ascertainment, using observed diagnostic and questionnaire evidence without phenotype reconstruction or auxiliary-trait integration. GWAS estimates were combined by inverse-variance meta-analysis. To assess alignment with the PGC depression target, we used LD-score regression (Bulik-Sullivan et al., 2015) and found a genetic correlation close to unity (unconstrained *r*^*_g_* = 1.011). Supplementary Note 1 specifies phenotype, harmonization and effective-sample-size rules.

### 4.3 Depression measurement and family validation

The MD reference phenotype used the LifetimeMDD definition (Cai et al., 2020; Dahl et al., 2023), which is based on UK Biobank’s mental-health questionnaire (Supplementary Note 2). Measured PGS performance depends on the depression definition and on the cohort in which it is evaluated: shallow definitions have shown lower SNP heritability and less depression-specific genetic signal than detailed diagnostic phenotypes (Cai et al., 2020; Dahl et al., 2023), and in the PGC’s own leave-one-cohort-out evaluation, the full PGC 2025 score (including 23andMe) explained 4.1% of liability variance in community-recruited cohorts but 7.0% in cohorts recruited from inpatient, outpatient or register settings (Adams et al., 2025, Table S6). Absolute estimates are therefore not directly comparable across definitions and cohorts, whereas the two scores were always compared on the same phenotype and participants.

We estimated population associations with the reference depression phenotype using logistic regression of case/control status on the full PGS, adjusting for sex, birth year, its square, assessment center, array and genetic principal components (PCs) 1–20 and obtained liability-scale *R*^2^ using established conversions assuming a population depression prevalence of 15% (see “Repeat measurement and liability” and Supplementary Note 2).

All SNIPar analyses (depression and adult outcomes) used sibships of the sibling validation sample, which excludes families with a genotyped parent, so both parental scores were imputed by SNIPar from the siblings’ genotypes (Young et al., 2022). Participant and parental scores were calculated using identical variants and SNP weights. We fitted two linear mixed models to the same participants, using the same covariates and score scaling:

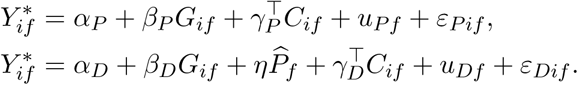

*Y ^∗^* denotes case/control status scaled to unit SD, *G* is the participant’s PGS, *P̂* is the (imputed) parental-score sum in the same units, *C* contains the covariates, and *u* denotes a family random intercept. The population coeffiicient *β_P_* estimates the association without parental-score adjustment, whereas *β_D_* estimates the association conditional on parental scores. The ratio *β_D_*/*β_P_*denotes the effect retained within families. Coeffiicient standard errors are SNIPar’s model-based estimates, and the interval for the ratio came from 4,000 whole-family bootstrap samples that refitted both models. As a robustness check, we also ran probit models, which gave nearly identical depression associations to the linear models on the probability scale (Supplementary Note 2).

Latent internalizing used 28 ordinal PHQ/GAD/neuroticism items in UK Biobank and 30 Youth Self-Report (YSR) items averaged across all available ABCD assessments (4.5-, 5.5-, 6- and 7-year follow-ups). Neuroticism used the 12-item Eysenck Personality Questionnaire–Revised Short Form. The hierarchies represented shared internalizing variation across depression, anxiety and neuroticism in UK Biobank, and anxious/depressed, withdrawn/depressed and somatic symptoms in ABCD. UK Biobank used weighted least squares (WLSMV) due to its ordinal item responses, with pairwise ordinal information. ABCD treated averaged item responses as continuous and used robust maximum likelihood with full-information handling of missing items (MLR/FIML) and family clustering. Sibling models included the family mean and the child’s deviation for each score, *G_if_* = *G̅_f_*+(*G_if_ G̅_f_*), in the latent regression, where the deviation coeffiicient estimated the within-family association. Within-family childhood phenome models included family fixed effects, with a separate intercept for each family. For UK Biobank anxiety, the estimated within-family advantage remained positive, but its confidence interval included zero when complete responses to all 28 questionnaire items were required (Supplementary Table S6B).

#### Repeat measurement and liability

Because case/control status indicates whether continuous liability exceeds a threshold, conversion to the liability scale depends on assumptions concerning prevalence and ascertainment. We transformed log-odds coeffiicients per score SD using standard conversions to the liability scale (Lee et al., 2012; Moore et al., 2025).

Recall or reporting error can further weaken the association between a case/control label and a PGS (Kendler et al., 1993). To correct for measurement error, we reconstructed the depression algorithm in two questionnaires administered in UK Biobank in 2016–2017 and 2022–2023. Of 79,072 participants for whom the reference phenotype could be constructed at both assessments, 25,022 (31.6%) changed classification after a median of 6.2 years (Supplementary Table S3C). We compared binary Cohen’s kappa, a three-state polychoric correlation and a two-component Gaussian-copula mixture as alternative measures of reliability *s*. The working correction divided the population log-odds coeffiicient by 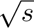 before applying the coeffiicient-derived liability mapping at assumed prevalence *K* = 0.15, following the PGC depression analysis (Adams et al., 2025). The uncorrected specification used *s* = 1. The polychoric specification is used in Figure 1A, and Table S3 compares alternatives.

The implied within-family liability-*R*^2^ in Figure 1A was the population summary multiplied by the squared same-score SNIPar direct/population ratio, assuming transfer to the full-score association in population variance units. Supplementary Note 2 gives the derivation, calibration and resampling details.

### 4.4 Adult health and disease associations

Twenty health, pain, cognitive and socioeconomic outcomes and 23 diagnoses were analyzed with and without parental PGS using SNIPar, with binary diagnoses treated as linear outcomes. Models adjusted for sex, birth year, its square and PCs 1–20. For sex-restricted cancers, covariates were birth year and PCs.

For adult outcomes, coeffiicient intervals used family-clustered robust (HC0) covariance, conditional on the fitted variance components, and direct/population ratio intervals used Fieller’s method (Fieller, 1954) with the same covariance. Fieller’s method inverts the test that the direct coeffiicient minus the ratio times the population coeffiicient equals zero. Unlike the delta method, it does not rely on a linear approximation and gives asymmetric or unbounded intervals when the denominator is imprecise. For each score, BH correction was applied to all 23 diagnoses, separately for population and direct-effect models. We used Fieller intervals also for portability reductions and observed-scale *R*^2^ gains, with jackknife or influence-function covariances, and log-ratio intervals for gains in latent coeffiicients (Supplementary Notes 2, 3 and 7). The core depression ratio instead came from the 4,000-sample whole-family bootstrap used for its liability-*R*^2^. The ratio and the model-implied within-family *R*^2^, which multiplies the population *R*^2^ by the squared ratio, therefore share one set of resampled families, which also re-estimates the reliability correction and the variance components. Throughout, confidence intervals are pointwise 95% intervals unless stated otherwise. In multiple-testing families with a fixed set of tests, a test that could not be computed was counted as *P* = 1, whereas the BH correction of household-moderation tests across outcomes and exposures included only the tests that could be computed.

Supplementary Note 4 gives the outcome definitions, measurement occasions, disease codes, control exclusions and model specifications.

### 4.5 Childhood phenome and developmental trajectories

We tested PGS associations with 2,328 ABCD outcomes using repeated observations across eligible assessment waves. We fitted linear models to continuous outcomes and to binary outcomes coded 0 or 1, adjusting for sex, baseline age, PCs 1–10, site and assessment wave. Standard errors accounted for correlations among observations from the same family.

We compared population and within-family associations in siblings and dizygotic twins. These models used identical participants and assessments, with the same PGS and outcome scaling. The within-family model included a separate intercept for each family, estimating associations from differences between siblings. For the 1,854 outcomes with estimates from the full population sample and both sibling models, we used regression slopes to summarize differences in association strength between models.

Slopes were constrained to pass through zero and corrected for sampling error. Confidence intervals used 10,000 draws, applying each family’s random weight consistently across outcomes and models to preserve their correlations (Supplementary Note 5).

Comparison models between our score and the PGC benchmarks used the same design and outcomes. We compared coeffiicient profiles and mean incremental observed-scale *R*^2^, defined relative to original outcome total variance, including for family fixed-effect models (Supplementary Note 5 and Tables S4 and S10). The population-profile correlation of 0.99 was estimated across all 2,328 outcomes. Family-model comparisons retained the 1,854-outcome common inventory.

#### Developmental trajectories

Six binary outcomes, selected as illustrative examples, used logistic models with assessment indicators and PGS-by-assessment interactions, adjusting for sex, baseline age, PCs 1–10, site and within-assessment age deviation. Probabilities were averaged over the empirical top and bottom score deciles using a common covariate distribution. Supplementary Note 5 and Table S11 give outcome definitions, recall periods and minimum participant/event counts.

### 4.6 Sex differences and gene-environment interaction

We fitted separate models in boys and girls, allowing covariate effects to differ by sex. Both sexes used the same reference PGS SDs and outcome SDs calculated across sexes. We tested differences between boys’ and girls’ coeffiicients across 726 psychiatric outcomes, accounting for relatedness between participants, including brothers and sisters. Positive differences indicate more positive coeffiicients in boys, which need not mean stronger associations.

We summarized the overall sex difference using a regression slope relating boys’ coeffiicients to girls’ coeffiicients, with the line constrained to pass through zero. Each outcome received equal weight, and the slope was corrected for sampling error and correlated estimation errors between sexes. Confidence intervals used 10,000 draws that reweighted families’ contributions to the estimates (Supplementary Note 6). This slope describes an average pattern, while individual outcomes can differ.

A sensitivity analysis standardized outcomes separately in boys and girls while keeping PGS scaling unchanged. Comparisons between the integrated and PGC scores used identical outcome sets, both within the psychiatric subset and across the full phenome.

#### Symptom content

For the exploratory content analysis, separate gpt-5.6-sol requests rated items from 1 (strongly boy-typical) to 9 (strongly girl-typical), using item wording or source descriptions, reporter and response options. The ratings describe whether symptoms are culturally perceived as more typical of boys or girls. They do not measure participants’ gender or how often symptoms actually occur. Requests contained no participant data, association results, previous ratings or conversation history. Ratings describe perceived cultural typicality among US adolescents aged 12–15. For 716 usable ratings, equally weighted item-level trends tested their relationship to coeffiicients in each sex and the between-sex trend difference.

Confidence intervals accounted for correlations across items and between relatives, including brothers and sisters. After standardizing outcomes separately in boys and girls, PGS associations in girls still increased with how girl-typical the symptoms were rated. However, the difference between girls’ and boys’ trends was attenuated. Supplementary Note 6 provides the methods, and the ratings are part of the Supplementary Data.

#### Gene-environment interaction

We examined baseline family resources (income and parental education), material hardship measured with seven items, and residential deprivation measured by the Area Deprivation Index (Kind and Buckingham, 2018). Higher resources indicate greater advantage, whereas higher hardship and deprivation indicate greater disadvantage. Exposures were assigned to families and standardized before selecting participants with outcome data. Families with missing required information or conflicting reports were excluded from that exposure’s analysis.

We tested whether PGS associations varied with each exposure using separate population models. Each model included the PGS, exposure and their interaction. Models also included a squared-exposure term and interactions of both the PGS and exposure with covariates (Keller, 2014). Latent internalizing models adjusted for sex, average age at YSR assessments, PCs 1–10 and site. Phenome models adjusted for sex, baseline age, PCs 1–10, site and assessment wave.

Analyses covered latent internalizing, the symptom subset and the full eligible phenome. Across outcomes, we summarized moderation using regression slopes relating interaction coeffiicients to PGS coeffiicients. The PGS coeffiicients represented associations at average exposure, averaged over the covariate values of the analysed observations. Slopes were corrected for sampling error and correlated errors between estimates.

For latent internalizing, sex differences were calculated as boys’ minus girls’ PGS coeffiicients, using the same latent scale for both sexes. For phenome summaries, we subtracted one from the boys-to-girls multiplier, so zero indicated no overall difference. Statistical tests and confidence intervals accounted for relatedness within families. Supplementary Note 6 gives the exposure definitions and multiple-testing corrections.

### 4.7 Population portability

UK Biobank used precomputed genetic-PC groups labeled United Kingdom, Nigeria, China and India, here called EUR, AFR, EAS and SAS. ABCD used parent-reported race/ethnicity groups: White (reference), Hispanic and Black (labelled African American). Multiple-group assignments were excluded except for the Hispanic group (Supplementary Note 1). The UK Biobank reference used the discovery-excluded family sample (United Kingdom PC group), and the AFR, EAS and SAS groups included all eligible scored participants, without a sibling requirement. Matched-score core comparisons estimated 100(1 *β_g_*/*β*_ref_) using group-specific PGS SDs and each score’s own reference-group denominator. Details are given in Supplementary Note 7.

## Acknowledgements

This research has been conducted using the UK Biobank Resource under Application Number #103244. This work uses data provided by patients and collected by the NHS as part of their care and support.

Furthermore, data used in the preparation of this article were obtained from the Adolescent Brain Cognitive Development™ (ABCD) Study (https://abcdstudy.org), held in the NIH Brain Development Cohorts Data Sharing Platform. This is a multisite, longitudinal study designed to recruit more than 10,000 children aged 9–10 and follow them over 10 years into early adulthood. The ABCD Study® is supported by the National Institutes of Health and additional federal partners under award numbers U01DA041048, U01DA050989, U01DA051016, U01DA041022, U01DA051018, U01DA051037, U01DA050987, U01DA041174, U01DA041106, U01DA041117, U01DA041028, U01DA041134, U01DA050988, U01DA051039, U01DA041156, U01DA041025, U01DA041120, U01DA051038, U01DA041148, U01DA041093, U01DA041089, U24DA041123, U24DA041147. A full list of supporters is available at https://abcdstudy.org/federal-partners.html. A listing of participating sites and a complete listing of the study investigators can be found at https://abcdstudy.org/consortium_members/. ABCD consortium investigators designed and implemented the study and/or provided data but did not necessarily participate in the analysis or writing of this report. This manuscript reflects the views of the authors and may not reflect the opinions or views of the NIH or ABCD consortium investigators. The ABCD data repository grows and changes over time. The ABCD data used in this report came from NBDC Release 7.0 (DOI https://doi.org/10.82525/8f3w-5260). DOIs can be found at https://www.nbdc-datahub.org/abcd-study.

We thank the participants and investigators of the All of Us Research Program and the NIH for making the research data available. We also acknowledge the participants and investigators of FinnGen.

## Ethics and consent

UK Biobank obtained approval from the North West Multi-centre Research Ethics Committee and informed consent from participants. ABCD obtained central or local institutional review board approval, parental consent and child assent. The All of Us Research Program obtained institutional review board approval and informed consent from participants. This study analyzed existing cohort data.

## Data availability

UK Biobank data are available to approved researchers through the UK Biobank Access Management System (https://www.ukbiobank.ac.uk/enable-your-research). ABCD data are available to approved researchers through the NIH Brain Development Cohorts Data Hub (https://www.nbdc-datahub.org/abcd-study). This study used the All of Us Research Program’s Controlled Tier v9 dataset, available to authorized researchers through the Researcher Workbench (https://www.researchallofus.org/data-tools/workbench/). Complete association results are in the Supplementary Data. Other study-level summary statistics are available from the authors upon reasonable request, subject to the cohorts’ data-use requirements.

## Code availability

Analysis code is available from the corresponding author on request.

## Use of AI tools

AI tools (OpenAI Codex and Anthropic Claude) assisted with writing and editing the manuscript and with compiling the Supplementary Information. The authors reviewed all content and take full responsibility for it.

## Footnotes

1 Following common usage, we use major depressive disorder (MDD) for the clinical diagnosis and major depression (MD) for the broader definitions available at biobank scale, which approximate it (McIntosh et al., 2019).

2 Each belonged to a sibship with at least two genotyped siblings, although most sibships contained only one participant with a depression classification (951 of 6,085 contained two or more). SNIPar imputes parental genotypes from all genotyped siblings, so every participant contributed. With the imputed parental score as a covariate, the direct coeffiicient reflects only the random inherited component of each participant’s score.

## References

ABCD Study. ABCD 7.0 data release: Release notes. https://docs.abcdstudy.org/v/7_0_2/documentation/release_notes/7_0.html, 2026. Release 13 May 2026; accessed 21 September 2026.

Mark J. Adams, Fabian Streit, Xiangrui Meng, Swapnil Awasthi, Brett N. Adey, Karmel W. Choi, et al. Trans-ancestry genome-wide study of depression identifies 697 associations implicating cell types and pharmacotherapies. Cell, 188(3):640–652.e9, 2025. doi: 10.1016/j.cell.2024.12.002. URL 10.1016/j.cell.2024.12.002.

Thomas D. Als, Mitja I. Kurki, Jakob Grove, Georgios Voloudakis, Karen Therrien, Elisa Tasanko, et al. Depression pathophysiology, risk prediction of recurrence and comorbid psychiatric disorders using genome-wide analyses. Nature Medicine, 29(7):1832–1844, 2023. doi: 10.1038/s41591-023-02352-1. URL 10.1038/s41591-023-02352-1.

A. Angold, E. J. Costello, A. Erkanli, and C. M. Worthman. Pubertal changes in hormone levels and depression in girls. Psychological Medicine, 29(5):1043–1053, 1999. doi: 10.1017/S0033291799008946. URL 10.1017/S0033291799008946.

Daniel J. Benjamin, David Cesarini, Patrick Turley, and Alexander Strudwick Young. Social-Science Genomics: Progress, Challenges, and Future Directions. Journal of Economic Literature, forthcoming. URL https://www.aeaweb.org/articles?id=10.1257/jel.20261604. Working-paper version: NBER Working Paper 32404, 2024. doi: 10.3386/w32404. URL 10.3386/w32404.

Yoav Benjamini and Yosef Hochberg. Controlling the False Discovery Rate: A Practical and Powerful Approach to Multiple Testing. Journal of the Royal Statistical Society Series B: Statistical Methodology, 57(1):289–300, 1995. doi: 10.1111/j.2517-6161.1995.tb02031.x. URL 10.1111/j.2517-6161.1995.tb02031.x.

Brendan Bulik-Sullivan, Hilary K. Finucane, Verneri Anttila, Alexander Gusev, Felix R. Day, Po-Ru Loh, et al. An atlas of genetic correlations across human diseases and traits. Nature Genetics, 47(11):1236–1241, 2015. doi: 10.1038/ng.3406.

Na Cai, Joana A. Revez, Mark J. Adams, Till F. M. Andlauer, Gerome Breen, Enda M. Byrne, et al. Minimal phenotyping yields genome-wide association signals of low specificity for major depression. Nature Genetics, 52(4):437–447, 2020. doi: 10.1038/s41588-020-0594-5. URL 10.1038/s41588-020-0594-5.

Claire E. Campbell, Jessica Morrel, W. James Gauderman, and Megan M. Herting. Effects of genetic risk for depression and household socioeconomic status on emotional behavior and brain development in early adolescence. Developmental Cognitive Neuroscience, 79:101692, 2026. doi: 10.1016/j.dcn.2026.101692. URL 10.1016/j.dcn.2026.101692.

Razieh Chegeni, Ragnhild Bang Nes, Rosa Cheesman, Ziada Ayorech, Eivind Ystrøm, and Espen Røysamb. Direct and indirect parental genetic effects on offspring susceptibility to internalizing problems across development. Nature Mental Health, 4(5):754–765, 2026. doi: 10.1038/s44220-026-00619-3. URL 10.1038/s44220-026-00619-3.

Andrew Dahl, Michael Thompson, Ulzee An, Morten Krebs, Vivek Appadurai, Richard Border, et al. Phenotype integration improves power and preserves specificity in biobank-based genetic studies of major depressive disorder. Nature Genetics, 55(12):2082–2093, 2023. doi: 10.1038/ s41588-023-01559-9. URL 10.1038/s41588-023-01559-9.

Yi Ding, Kangcheng Hou, Ziqi Xu, Aditya Pimplaskar, Ella Petter, Kristin Boulier, et al. Polygenic scoring accuracy varies across the genetic ancestry continuum. Nature, 618 (7966):774–781, 2023. doi: 10.1038/s41586-023-06079-4. URL doi.org/10.1038/s41586-023-06079-4.

E. C. Fieller. Some Problems in Interval Estimation. Journal of the Royal Statistical Society Series B: Statistical Methodology, 16(2):175–185, 1954. doi: 10.1111/j.2517-6161.1954.tb00159.x. URL 10.1111/j.2517-6161.1954.tb00159.x.

E. I. Fried, R. M. Nesse, C. Guille, and S. Sen. The differential influence of life stress on individual symptoms of depression. Acta Psychiatrica Scandinavica, 131(6):465–471, 2015. doi: 10.1111/ acps.12395. URL 10.1111/acps.12395.

Eiko I. Fried and Randolph M. Nesse. Depression is not a consistent syndrome: An investigation of unique symptom patterns in the STAR*D study. Journal of Affective Disorders, 172:96–102, 2015. doi: 10.1016/j.jad.2014.10.010. URL 10.1016/j.jad.2014.10.010.

Poppy Z. Grimes, Mark J. Adams, Gladi Thng, Amelia J. Edmonson-Stait, Yi Lu, Andrew McIntosh, et al. Genetic Architectures of Adolescent Depression Trajectories in 2 Longitudinal Population Cohorts. JAMA Psychiatry, 81(8):807–816, 2024. doi: 10.1001/jamapsychiatry. 2024.0983. URL 10.1001/jamapsychiatry.2024.0983.

Andrew D. Grotzinger, Josefin Werme, Wouter J. Peyrot, Oleksandr Frei, Christiaan de Leeuw, Lucy K. Bicks, et al. Mapping the genetic landscape across 14 psychiatric disorders. Nature, 649(8096):406–415, 2026. doi: 10.1038/s41586-025-09820-3. URL 10.1038/s41586-025-09820-3.

Junming Guan, Tammy Tan, Seyed Moeen Nehzati, Michael Bennett, Patrick Turley, Daniel J. Benjamin, and Alexander Strudwick Young. Family-based genome-wide association study designs for increased power and robustness. Nature Genetics, 57(4):1044–1052, 2025. doi: 10.1038/s41588-025-02118-0. URL 10.1038/s41588-025-02118-0.

Sture Holm. A simple sequentially rejective multiple test procedure. Scandinavian Journal of Statistics, 6(2):65–70, 1979. URL https://www.jstor.org/stable/4615733.

Laurence J. Howe, Michel G. Nivard, Tim T. Morris, Ailin F. Hansen, Humaira Rasheed, Yoonsu Cho, et al. Within-sibship genome-wide association analyses decrease bias in estimates of direct genetic effects. Nature Genetics, 54(5):581–592, 2022. doi: 10.1038/s41588-022-01062-7. URL 10.1038/s41588-022-01062-7.

Floris Huider, Yuri Milaneschi, René Pool, Bernardo de A. P. C. Maciel, Scott D. Gordon, Jihee Han, et al. Genetics of major depressive disorder in a homogeneous population with uniform phenotyping. Molecular Psychiatry, 31(10):5889–5899, 2026. doi: 10.1038/s41380-026-03666-5. URL 10.1038/s41380-026-03666-5.

S. C. Kanjira, M. J. Adams, Y. Jiang, C. Tian, 23andMe Research Team, C. M. Lewis, et al. Polygenic prediction of major depressive disorder and related traits in African ancestries UK Biobank participants. Molecular Psychiatry, 30(1):151–157, 2025. doi: 10.1038/ s41380-024-02662-x. URL 10.1038/s41380-024-02662-x.

Matthew C. Keller. Gene × Environment Interaction Studies Have Not Properly Controlled for Potential Confounders: The Problem and the (Simple) Solution. Biological Psychiatry, 75(1):18–24, 2014. doi: 10.1016/j.biopsych.2013.09.006. URL 10.1016/j.biopsych.2013.09.006.

Kenneth S. Kendler, Michael C. Neale, Ronald C. Kessler, Andrew C. Heath, and Lindon J. Eaves. The lifetime history of major depression in women: Reliability of diagnosis and heritability. Archives of General Psychiatry, 50(11):863–870, 1993. doi: 10.1001/archpsyc.1993.01820230054003. URL 10.1001/archpsyc.1993.01820230054003.

Amy J.H. Kind and William R. Buckingham. Making Neighborhood-Disadvantage Metrics Accessible — The Neighborhood Atlas. New England Journal of Medicine, 378(26):2456–2458, 2018. doi: 10.1056/NEJMp1802313. URL 10.1056/NEJMp1802313.

Sang Hong Lee, Michael E. Goddard, Naomi R. Wray, and Peter M. Visscher. A Better Coeffiicient of Determination for Genetic Profile Analysis. Genetic Epidemiology, 36(3):214–224, 2012. doi: 10.1002/gepi.21614. URL 10.1002/gepi.21614.

Luke R. Lloyd-Jones, Jian Zeng, Julia Sidorenko, Loïc Yengo, Gerhard Moser, Kathryn E. Kemper, et al. Improved polygenic prediction by Bayesian multiple regression on summary statistics. Nature Communications, 10(1):5086, 2019. doi: 10.1038/s41467-019-12653-0. URL 10.1038/s41467-019-12653-0.

Ani Manichaikul, Josyf C. Mychaleckyj, Stephen S. Rich, Kathy Daly, Michèle M. Sale, and WeiMin Chen. Robust relationship inference in genome-wide association studies. Bioinformatics, 26 (22):2867–2873, 2010. doi: 10.1093/bioinformatics/btq559. URL 10.1093/ bioinformatics/btq559.

Andrew M. McIntosh, Patrick F. Sullivan, and Cathryn M. Lewis. Uncovering the Genetic Architecture of Major Depression. Neuron, 102(1):91–103, 2019. doi: 10.1016/j.neuron.2019.03.022. URL 10.1016/j.neuron.2019.03.022.

T. E. Moffiitt, A. Caspi, A. Taylor, J. Kokaua, B. J. Milne, G. Polanczyk, et al. How common are common mental disorders? Evidence that lifetime prevalence rates are doubled by prospective versus retrospective ascertainment. Psychological Medicine, 40(6):899–909, 2010. doi: 10.1017/S0033291709991036. URL 10.1017/S0033291709991036.

Spencer Moore, Ivan Davidson, Jonathan Anomaly, Jeremiah H. Li, Mohammad Ahangari, Lauren Moissiy, Michael Christensen, Alexander Strudwick Young, David Stern, and Tobias Wolfram. Development and validation of polygenic scores for within-family prediction of disease risks. medRxiv, 2025. doi: 10.1101/2025.08.06.25333145. URL https://www.medrxiv.org/content/10.1101/2025.08.06.25333145v2. Version 2, posted 14 August 2025.

Thuy-Dung Nguyen, Arvid Harder, Ying Xiong, Kaarina Kowalec, Sara Hägg, Na Cai, et al. Genetic heterogeneity and subtypes of major depression. Molecular Psychiatry, 27 (3):1667–1675, 2022. doi: 10.1038/s41380-021-01413-6. URL 10.1038/s41380-021-01413-6.

Sarah E. Paul, Sarah M. C. Colbert, Aaron J. Gorelik, Emma C. Johnson, Alexander S. Hatoum, David A. A. Baranger, et al. A phenome-wide association study of cross-disorder genetic liability in youth genetically similar to individuals from European reference populations. Nature Mental Health, 2(11):1327–1341, 2024. doi: 10.1038/s44220-024-00313-2. URL 10.1038/s44220-024-00313-2.

Florian Privé, Hugues Aschard, Shai Carmi, Lasse Folkersen, Clive Hoggart, Paul F. O’Reilly, et al. Portability of 245 polygenic scores when derived from the UK Biobank and applied to 9 ancestry groups from the same cohort. The American Journal of Human Genetics, 109(1): 12–23, 2022. doi: 10.1016/j.ajhg.2021.11.008. URL 10.1016/j.ajhg.2021.11.008.

Jian Rong, Xueqin Wang, Pan Cheng, Dan Li, and Dahai Zhao. Global, regional and national burden of depressive disorders and attributable risk factors, from 1990 to 2021: results from the 2021 Global Burden of Disease study. The British Journal of Psychiatry, 227(4):688–697, 2025. doi: 10.1192/bjp.2024.266. URL 10.1192/bjp.2024.266.

Rachel H. Salk, Janet S. Hyde, and Lyn Y. Abramson. Gender differences in depression in representative national samples: Meta-analyses of diagnoses and symptoms. Psychological Bulletin, 143(8):783–822, 2017. doi: 10.1037/bul0000102. URL 10.1037/bul0000102.

Tabea Schoeler, Doug Speed, Eleonora Porcu, Nicola Pirastu, Jean-Baptiste Pingault, and Zoltán Kutalik. Participation bias in the UK Biobank distorts genetic associations and downstream analyses. Nature Human Behaviour, 7(7):1216–1227, 2023. doi: 10.1038/s41562-023-01579-9. URL 10.1038/s41562-023-01579-9.

Xueyi Shen, David M. Howard, Mark J. Adams, W. David Hill, Toni-Kim Clarke, Major Depressive Disorder Working Group of the Psychiatric Genomics Consortium, et al. A phenome-wide association and Mendelian Randomisation study of polygenic risk for depression in UK Biobank. Nature Communications, 11(1):2301, 2020. doi: 10.1038/s41467-020-16022-0. URL 10.1038/s41467-020-16022-0.

John R. Shorter, Joëlle A. Pasman, Siim Kurvits, Andreas Jangmo, Joonas Naamanka, Arvid Harder, et al. Genome-wide association analyses identify distinct genetic architectures for early-onset and late-onset depression. Nature Genetics, 57(12):2972–2979, 2025. doi: 10.1038/ s41588-025-02396-8. URL 10.1038/s41588-025-02396-8.

Cathie Sudlow, John Gallacher, Naomi Allen, Valerie Beral, Paul Burton, John Danesh, et al. UK Biobank: An Open Access Resource for Identifying the Causes of a Wide Range of Complex Diseases of Middle and Old Age. PLOS Medicine, 12(3):e1001779, 2015. doi: 10.1371/journal. pmed.1001779. URL 10.1371/journal.pmed.1001779.

Patrick F. Sullivan, Michael C. Neale, and Kenneth S. Kendler. Genetic Epidemiology of Major Depression: Review and Meta-Analysis. American Journal of Psychiatry, 157(10):1552–1562, 2000. doi: 10.1176/appi.ajp.157.10.1552. URL 10.1176/appi.ajp.157. 10.1552.

Tammy Tan, Hariharan Jayashankar, Junming Guan, Seyed Moeen Nehzati, Mahdi Mir, Michael Bennett, et al. Family-GWAS reveals effects of environment and mating on genetic associations. medRxiv, 2024. doi: 10.1101/2024.10.01.24314703. URL 10.1101/2024.10.01.24314703. Preprint.

Jodi T. Thomas, Jackson G. Thorp, Floris Huider, Poppy Z. Grimes, Rujia Wang, Pierre Youssef, et al. Sex-stratified genome-wide association meta-analysis of major depressive disorder. Nature Communications, 16(1):7960, 2025. doi: 10.1038/s41467-025-63236-1. URL 10.1038/s41467-025-63236-1.

Jackson G. Thorp, Adrian I. Campos, Andrew D. Grotzinger, Zachary F. Gerring, Jiyuan An, Jue-Sheng Ong, et al. Symptom-level modelling unravels the shared genetic architecture of anxiety and depression. Nature Human Behaviour, 5(10):1432–1442, 2021. doi: 10.1038/ s41562-021-01094-9. URL 10.1038/s41562-021-01094-9.

Victória Trindade Pons, Albertine J. Oldehinkel, and Hanna M. van Loo. Genetic nurture effects in depressive and anxiety disorders and symptoms, and in related traits. Molecular Psychiatry, 30(12):5694–5700, 2025. doi: 10.1038/s41380-025-03265-w. URL 10.1038/s41380-025-03265-w.

Patrick Turley, Raymond K. Walters, Omeed Maghzian, Aysu Okbay, James J. Lee, Mark Alan Fontana, et al. Multi-trait analysis of genome-wide association summary statistics using MTAG. Nature Genetics, 50(2):229–237, 2018. doi: 10.1038/s41588-017-0009-4. URL 10.1038/s41588-017-0009-4.

Jessica Tyrrell, Jie Zheng, Robin Beaumont, Kathryn Hinton, Tom G. Richardson, Andrew R. Wood, et al. Genetic predictors of participation in optional components of UK Biobank. Nature Communications, 12(1):886, 2021. doi: 10.1038/s41467-021-21073-y. URL 10.1038/s41467-021-21073-y.

Carl Veller and Graham M. Coop. Interpreting population- and family-based genome-wide association studies in the presence of confounding. PLOS Biology, 22(4):e3002511, 2024. doi: 10.1371/journal.pbio.3002511. URL 10.1371/journal.pbio.3002511.

Ying Wang, Jing Guo, Guiyan Ni, Jian Yang, Peter M. Visscher, and Loic Yengo. Theoretical and empirical quantification of the accuracy of polygenic scores in ancestry divergent populations. Nature Communications, 11(1):3865, 2020. doi: 10.1038/s41467-020-17719-y. URL 10.1038/s41467-020-17719-y.

Monika A. Waszczuk, Jiaju Miao, Anna R. Docherty, Andrey A. Shabalin, Katherine G. Jonas, Giorgia Michelini, et al. General v. specific vulnerabilities: polygenic risk scores and higher-order psychopathology dimensions in the Adolescent Brain Cognitive Development (ABCD) Study. Psychological Medicine, 53(5):1937–1946, 2023. doi: 10.1017/S0033291721003639. URL 10.1017/S0033291721003639.

Tobias Wolfram, Spencer Moore, Jeremiah H. Li, Jonathan Anomaly, Ivan Davidson, and Michael Christensen. Interpreting Polygenic Prediction of Cognitive Ability: Evidence for Direct, Reliable, and Portable Genetic Effects. Intelligence & Cognitive Abilities, 2(1):1–19, 2026. doi: 10.65550/001c.158459. URL 10.65550/001c.158459.

Alexander I. Young, Seyed Moeen Nehzati, Stefania Benonisdottir, Aysu Okbay, Hariharan Jayashankar, Chanwook Lee, et al. Mendelian imputation of parental genotypes improves estimates of direct genetic effects. Nature Genetics, 54(6):897–905, 2022. doi: 10.1038/ s41588-022-01085-0. URL 10.1038/s41588-022-01085-0.

Zhili Zheng, Shouye Liu, Julia Sidorenko, Ying Wang, Tian Lin, Loic Yengo, et al. Leveraging functional genomic annotations and genome coverage to improve polygenic prediction of complex traits within and between ancestries. Nature Genetics, 56(5):767–777, 2024. doi: 10.1038/s41588-024-01704-y. URL 10.1038/s41588-024-01704-y.

