## Supplementary Information for "Polygenic Prediction of Depression Liability Reveals Broad Genetic Associations Across Development, Families, and Populations"

This Supplementary Information describes the samples, measures and statistical models behind each result in the main text and reports additional results. Supplementary Note 1 covers the samples, genotype processing and polygenic scores used throughout. Supplementary Notes 2 to 7 follow the order of the main text. Complete association results for all outcomes are provided in the Supplementary Data workbook.

Throughout, "integrated score" refers to our depression polygenic score and "PGC score" to the benchmark score built from Psychiatric Genomics Consortium summary statistics. The two scores were always compared on identical participants, outcomes and covariates. UK Biobank population groups are labelled EUR (reference), AFR, EAS and SAS, and ABCD groups are labelled White (reference), Hispanic and African American (Supplementary Notes 1 and 7).

### Supplementary Note 1: Samples, genotypes and polygenic scores

This note describes the validation samples, genotype processing, score standardization and the construction of the PGC benchmark scores. These elements are shared by all later analyses.

**UK Biobank.** We set aside for validation all genetically inferred full-sibling and parent–offspring pedigrees among participants in the United Kingdom genetic-ancestry group defined by Privé et al. (2022), and excluded every pedigree member from local discovery. Pedigrees were built using kinship, age and sex. The sibling validation sample comprised the reserve's sibships with no genotyped parent. Participants flagged as heterozygosity or missingness outliers (field 22027) or as having a sex-chromosome aneuploidy (field 22019) were excluded.

The depression analyses (Supplementary Note 2) additionally required British self-reported ethnicity (field 21000), the UK Biobank genetic ethnic grouping "Caucasian" (field 22006) and agreement between reported and genetic sex (fields 31 and 22001). Each participant needed a case or control classification and complete genotype and covariate data, but their siblings did not need to be classified. The latent internalizing analysis (Supplementary Note 3) instead required at least two siblings per family who had each answered at least one symptom item and had complete covariates.

**ABCD.** We used phenotypes from ABCD Release 7 (ABCD Study, 2026), genotypes imputed by the consortium, and the cohort's family and demographic records (family identifiers, parent-reported race and ethnicity, sex and genetic principal components). Family analyses used non-twin siblings and dizygotic twins. Monozygotic twins and twins with missing or conflicting zygosity were excluded. A family entered the reference sample if at least two of its members had complete scores and covariates and enough internalizing data: at least two observed internalizing composites among the parent-reported Child Behavior Checklist (CBCL), the Youth Self-Report (YSR) and the youth Brief Problem Monitor (BPM), or one such composite plus at least three teacher-reported BPM items. The latent YSR analysis then required at least one answered YSR item per child, but not a second YSR-reporting sibling (Table S1). Broader population analyses used all eligible White children, including singletons, siblings and twins. Sex was the parent-reported sex at baseline, or the sex assigned at birth in the Release 7 participant table for the latent family analysis.

ABCD population groups were based on parent-reported race and ethnicity (Wolfram et al., 2026). Children reported as Hispanic formed the Hispanic group, whatever their reported race. Each remaining child was assigned to a group if exactly one of four categories applied: White, Black, Asian Indian, or East Asian (Chinese, Filipino, Japanese or Korean). We analysed the White (reference), Hispanic and Black (labelled African American) groups. The East Asian and Asian Indian groups were below the 150-child minimum for model fitting (Supplementary Note 7). Children reported in more than one of the four categories were excluded, and so were children reported in none of them.

Table S1: ABCD reference and YSR analysis sample flow

| Selection | Children | Families |
|---|---|---|
| White genotyped children with both scores and complete reference covariates | 6,325 | 5,129 |
| Eligible non-twin siblings or confirmed dizygotic twins | 1,398 | 761 |
| Meeting the multi-instrument availability requirement | 1,396 | 760 |
| Reference sample: at least two eligible relatives per family | 1,268 | 632 |
| YSR analysis: at least one observed YSR item | 1,172 | 593 |

Each row applies an additional restriction. Score moments and family means used the reference sample before YSR restriction. The final analysis included 17 families with only one YSR-observed child. Their family means included the other reference-eligible relatives.

**Genotypes and scoring.** In UK Biobank, imputed genotypes were restricted to biallelic variants with at most 5% missing calls, minor-allele frequency of at least 1% and Hardy–Weinberg $P \geq 10^{-50}$. Participants with more than 3% missing genotypes (10% in the African-ancestry groups) were removed. Full scores were computed with PLINK 2 (Chang et al., 2015) from imputed dosages, as the average allele-weighted dosage over the scored variants. Weight variants that were absent from the target data or had incompatible alleles were skipped, and missing genotypes were replaced by their expected dosage given the allele frequency.

Scores for the family analyses were computed by SNIPar (Young et al., 2022). SNIPar used hard-called genotypes, converted from the imputed data for the family reserve after restricting to a list of well-imputed variants (imputation INFO $> 0.99$) and removing variants with Hardy–Weinberg $P < 10^{-6}$. Parental genotypes were imputed from the genotypes of siblings and the genome segments they share. Full-sibling and parent–offspring relationships among UK-ancestry participants were inferred with KING 2.3.2 (Manichaikul et al., 2010), and SNIPar built pedigrees using year of birth and sex. Identity-by-descent segments were inferred with SNIPar's `ibd.py` from directly genotyped array SNPs (607,402 of 784,256 retained), using the HapMap II genetic map and a genotyping-error probability of $10^{-4}$. Child and parental scores were computed from the same variants and weights: those present in the child's genotypes, in the imputed parental genotypes and among the score's non-zero weights. The within-family analyses of depression (Supplementary Note 2) therefore used this parental-compatible version of each score, while the logistic model of depression used the full score.

In ABCD, scores were computed from hard calls derived from the consortium's imputed genotypes.

**Score standardization.** Scores were standardized to unit variance in the study sample of each analysis before restricting to participants with a particular outcome (for adult UK Biobank outcomes, within each outcome's sample, which changed the score SD by less than 1%). There were two exceptions. The UK Biobank depression portability models standardized each group's scores among its participants with a depression classification and complete covariates. The ABCD latent population, moderation and portability models standardized each group's scores among children with at least one answered YSR item (Supplementary Notes 6 and 7).

**PGC benchmark scores.** In ABCD, the benchmark was an SBayesR score (Lloyd-Jones et al., 2019) computed from the published European PGC MDD2025 summary statistics without 23andMe (Adams et al., 2025), which include UK Biobank. Because those statistics include UK Biobank, the UK Biobank benchmark was rebuilt. The PGC statistics without UK Biobank were meta-analysed with a local UK Biobank depression GWAS that excluded every validation participant and their relatives (kinship $\geq 0.0442$).

The local GWAS phenotype used observed evidence:

- *Cases* met a detailed lifetime definition based on the Composite International Diagnostic Interview (CIDI), reported depression, reported a professional diagnosis, or had a hospital diagnosis of F32 or F33.

- *Exclusions* took priority: bipolar, psychotic or personality disorders.
- *Controls* required observed negative evidence and no ambiguous evidence. Ambiguous evidence was antidepressant use without a qualifying diagnosis, positive core depressive symptoms, probable depression, or another mood-disorder hospital code.

Unrelated participants were then selected without regard to case status. Associations were tested with GCTA fastGWA (Jiang et al., 2019), using a linear model of the 0/1 phenotype on hard-called genotypes, with age, sex, genotyping array and principal components 1–20 as covariates. Variants required a minor-allele frequency above 0.01, at least 20 copies of the minor allele and genotype missingness below 0.02.

Linear coefficients and standard errors of the local GWAS were divided by $k(1-k)$, where $k$ is its case fraction, to approximate log odds ratios. The local and PGC results were then meta-analysed by allele-aligned inverse-variance weighting. A variant kept its PGC estimate alone if it had no usable local result, if its allele frequencies differed by more than 0.10 between the two sources, or if it was strand-ambiguous with either frequency within 0.10 of 0.5. The effective sample size was the PGC value plus $4/(1/N_{\text{cases}} + 1/N_{\text{controls}})$ for the local GWAS. Both benchmark scores were estimated with SBayesR using the European banded HapMap3 linkage-disequilibrium reference.

**Discovery-signal comparison.** The integrated GWAS (final multi-trait statistics, before effect estimation) was compared with the published PGC GWAS without 23andMe on 960,778 autosomal HapMap3 variants that were also present in the UK Biobank focal depression GWAS. The MHC region, variants with allele frequency outside 0.01–0.99, strand-ambiguous variants and variants with $\chi^2 \geq 80$ in either GWAS were excluded. One cross-trait LD-score regression (Bulik-Sullivan et al., 2015) with 1000 Genomes European LD scores and free intercepts was fitted with the same fixed sample size for both GWASs, so that the ratio of the two heritability estimates equals the ratio of LD-score slopes ($N \times h^2_{\text{SNP}}$). The 95% interval of the ratio was $\exp(\log r \pm 1.96\,\text{SE})$, with the SE from a paired 200-block jackknife of the log ratio. Uniformity was checked with the ratio of mean $\chi^2$ excess over the intercepts within LD-score quintiles, and the genetic correlation was the unconstrained LD-score estimate with its block-jackknife SE.

**Similarity of the two benchmark GWASs.** Cross-trait LD-score regression (Bulik-Sullivan et al., 2015) compared the published PGC GWAS with our PGC-plus-UK Biobank GWAS. The unconstrained estimate of the genetic correlation on common HapMap sites was 1.011 (SE 0.00115, 95% interval 1.009–1.014), showing that the two sets of summary statistics are nearly identical.

### Supplementary Note 2: Depression in UK Biobank and ABCD

**Case and control definition.** We used the LifetimeMDD definition (Cai et al., 2020; Dahl et al., 2023) (Table S2): Cases required at least two weeks of depressed mood or loss of interest, at least five of eight lifetime symptom domains and substantial interference with normal roles, or alternatively at least five of the nine current symptoms present nearly every day, including depressed mood or loss of interest, and the same impairment. As in the original definition (Cai et al., 2020), cases with a self-reported psychotic, bipolar or substance-use disorder were left unclassified, which implements DSM-5 criteria C–E for cases. Controls explicitly denied a period of prolonged depressed mood and did not meet the case rule, while participants who met neither definition were also left unclassified. Cai et al. (2020) describe the controls only as participants without the case criteria. Applied to the same white British quality-controlled sample, our implementation gave 16,189 cases and 49,846 controls (published: 16,301 and 50,870), case counts within 14 of the published counts at every assessment centre, and the published lead association (rs926552, odds ratio 0.902 versus 0.903). In the analysis sample below, 1,637 of the 1,679 cases qualified only through the lifetime route, 41 through both routes and one only through the current-symptom route.

**Population and within-family models.** The analysis sample comprised 7,054 participants (1,679 cases) from 6,085 families of the sibling validation sample (Supplementary Note 1). All models adjusted for sex, birth year and its square, assessment centre, genotyping array and principal components 1–20. The population association of the full score was estimated by logistic regression. Within-family associations were estimated with SNIPar (Young et al., 2022) using the parental-compatible score. SNIPar fits two linear mixed models with family random intercepts by restricted maximum likelihood: a child-only model, which estimates the population association, and a model that adds the sum of the two imputed parental scores, which estimates the direct (within-family) association. In these linear models the binary outcome was standardized. Chromosome scores were combined before SNIPar's genome-wide adjustment for assortative mating. SNIPar applies this adjustment only when the estimated correlation between the two parents' scores is large relative to its standard error. The adjustment was therefore applied to the integrated score (estimated correlation 0.018, SE 0.008) but not to the PGC score, whose estimated correlation was too small (0.008, SE 0.008). Omitting the adjustment changed the integrated score's direct/population ratio by less than 0.001. Coefficients were estimated by generalized least squares at the REML variance components, and we report SNIPar's model-based standard errors. Because each imputed parental score is the expected score given the siblings' genotypes and their identity-by-descent sharing, its imputation error enters the residual, so estimates and standard errors remain valid without further correction (Young et al., 2022). The interval for the ratio came from the whole-family bootstrap described below. The ratio of the direct to the population coefficient, $r = \widehat{\beta}_D/\widehat{\beta}_P$, summarizes how much of the population association is retained within families.

**Stability of the depression classification.** Reporting error in a single case–control classification attenuates associations with a polygenic score. To estimate how stable the classification is, we applied the same rules to the matching items of the first (2016–2017) and repeat (2022–2023) mental-health questionnaires. At each assessment a respondent was a control, a case, or unclassified. The unclassified group reported qualifying symptoms without meeting the case or control definition, missing answers alone did not place anyone in it, and excluded cases were removed. We used all 79,072 respondents with an interpretable state at both assessments, including people outside the validation sample, as long as they reported British ethnicity, were in the genetic ethnic grouping labeled "Caucasian", passed the sex and aneuploidy checks of the depression sample and were in UK Biobank's unrelated set used for genetic principal components (field 22020). Table S3C shows the transitions.

We summarized stability in three ways (Table S3D):

- *Binary kappa.* Cohen's kappa (Cohen, 1960), $\kappa = (p_o - p_e)/(1 - p_e)$, among respondents who were a case or a control at both assessments.
- *Polychoric correlation.* A three-state polychoric correlation (Olsson, 1979), which assumes one bivariate normal liability and was estimated by maximum likelihood with separate thresholds for each questionnaire.
- *Stability mixture.* A two-component Gaussian-copula mixture. It fixes the thresholds at the observed marginal proportions and estimates two ordered correlations and a mixing weight $\pi$:

  $$p_{ab} = \pi p_{ab}(\rho_{\text{low}}) + (1-\pi)p_{ab}(\rho_{\text{high}}), \qquad s = \pi\rho_{\text{low}} + (1-\pi)\rho_{\text{high}}.$$

  The two components describe the response distribution, not clinical subgroups.

Using a stability estimate $s$ as a reliability assumes a stable standardized liability $T$, measured by two equally reliable questionnaires whose errors are independent of each other, of $T$ and of the score: $U_j = \sqrt{s}\,T + \sqrt{1-s}\,\varepsilon_j$. Associations are then attenuated by $\sqrt{s}$, and we corrected the log odds ratio as $b/\sqrt{s}$, which is an approximation for covariate-adjusted logistic coefficients. The mixture reproduced the transition table more closely than the polychoric model: the largest difference between observed and fitted cell proportions was 0.62 versus 2.76 percentage points. Neither fit statistic accounts for family structure, and a closer fit does not by itself validate the reliability interpretation. Figure 1A uses the polychoric correlation, whereas the mixture implies a slightly larger correction (Table S3A).

**Liability-scale variance explained.** We converted the full-score log odds ratio into approximate liability-scale $R^2$ with the conversion in Supplementary Table 5 of Moore et al. (2025), which includes the ascertainment adjustment of Lee et al. (2012). Let $b$ be the log odds ratio per score SD, $\widetilde{b} = b/\sqrt{s}$ its corrected value ($s = 1$ without correction), $P$ the case fraction of the analysis sample and $K = 0.15$ the assumed population prevalence. Under a binormal model, in which scores are normally distributed with equal variance in cases and controls, the standardized case–control difference in the score, $d$, satisfies

$$\widetilde{b} = d\sqrt{1 + d^2P(1-P)}, \qquad R_o^2 = \frac{d^2P(1-P)}{1 + d^2P(1-P)}.$$

With $a = \widetilde{b}^2P(1-P)$, the observed-scale value is

$$R_o^2 = \frac{4a}{\left(1 + \sqrt{1+4a}\right)^2}.$$

With $t = \Phi^{-1}(1-K)$, $z = \phi(t)$ and $i = z/K$, define

$$C = \frac{[K(1-K)]^2}{z^2P(1-P)}, \qquad \theta = i\frac{P-K}{1-K}\left(i\frac{P-K}{1-K} - t\right).$$

The population and model-implied within-family liability summaries are then

$$R^2_{\text{pop}}(s) = \frac{CR_o^2}{1 + C\theta R_o^2}, \qquad R^2_{\text{WF,implied}}(s) = R^2_{\text{pop}}(s)\, r^2.$$

These summaries are derived from association coefficients under a binormal approximation and an assumed prevalence. Sex-specific values (Results) refitted the logistic model separately in women and men, with a sex-specific repeat-stability correction and the same $K = 0.15$ in both sexes.

**Uncertainty.** Confidence intervals came from 4,000 whole-family bootstrap samples, stratified by membership of the validation and calibration samples. Repeat respondents without relatives in the reserve were resampled as single-person units. The same bootstrap weights were used for both scores and all specifications, so paired differences account for the overlap between samples. In each bootstrap sample we refitted the logistic models and the four SNIPar models (two per score), recalculated the transition statistics and the mixture thresholds, and updated the case fraction and the ascertainment terms $C$ and $\theta$. Score references and $K$ were held fixed. Intervals are bootstrap percentiles, and ratios above one and $R^2$ values were not truncated.

**Observed-scale variance explained.** The main analyses compare the scores on different scales: liability $R^2$ for depression, latent coefficients for internalizing and coefficient profiles for the phenome. To compare them on one scale, we computed the incremental variance explained on the observed scale, $(\text{SSE}_0 - \text{SSE}_1)/\text{TSS}$, where $\text{SSE}_0$ and $\text{SSE}_1$ are the residual sums of squares without and with the score and TSS is the total sum of squares. Binary outcomes were analysed as 0/1 variables without correction for measurement error. Both scores used identical rows and covariates. The UK Biobank internalizing outcome was the mean of the raw PHQ, GAD and neuroticism sums among participants who answered all 28 items, and the anxiety and neuroticism outcomes were the corresponding scale sums. The ABCD outcome was the mean of the three YSR scale sums, averaged over visits with complete answers. The adult panel averages all 43 UK Biobank life, health and disease outcomes with equal weight, without selection on significance. Uncertainty came from 40 whole-family deletions shared by both scores, refitting every model and variance, with paired $t_{39}$ Fieller intervals. A deletion in which an outcome was constant contributed zero for both scores. Table S4 gives the results.

**Probit models.** The population and within-family models of binary outcomes are linear. As a check, we refitted the child-only and parental-adjusted models with a probit link and a family random intercept,

$$\Pr(Y_{if} = 1 \mid G_{if}, P_f, C_{if}, u_f) = \Phi(\alpha + \beta G_{if} + \eta P_f + \gamma^\top C_{if} + u_f), \qquad u_f \sim N(0, \tau^2),$$

with $\eta = 0$ in the child-only model, for core depression, the 23 diagnoses and the seven binary pain outcomes. Participants, covariates and score units were identical to the linear models. We compared the linear coefficients, expressed as changes in probability, with the probit average marginal effects integrated over the family intercept,

$$\text{AME} = \frac{1}{n}\sum_i \frac{\beta}{\sqrt{1+\tau^2}}\, \phi\!\left(\frac{a_i}{\sqrt{1+\tau^2}}\right),$$

where $a_i$ is the linear predictor without the random intercept. Multiplied by 100, both are percentage points per score SD. For depression, intervals for the estimates, the link differences and the parental-adjustment comparisons came from 200 paired whole-family bootstrap samples that refitted both links, both scores and both parental specifications. For the adult outcomes, probit intervals used the delta method, conditional on the fitted random-intercept variance and the observed covariate distribution, and the linear intervals used the family-clustered covariance. These comparisons are descriptive, without paired tests.

In the depression sample (7,054 participants, 1,679 cases, 6,085 families), the two links agreed closely. For the integrated score, the child-only association was 7.499 percentage points per SD in the linear model (95% CI 6.673 to 8.438) and 7.453 (6.637 to 8.312) in the probit model, a difference of −0.046 (−0.177 to 0.067). After parental adjustment it was 7.737 (5.740 to 9.489) and 7.709 (5.785 to 9.374). For the PGC score, the corresponding values were 6.113 (5.194 to 7.085) and 6.107 (5.261 to 7.065) without, and 6.254 (4.782 to 8.144) and 6.273 (4.744 to 8.026) with, parental adjustment. Across the 30 adult binary outcomes, both scores and both specifications, the probit-minus-linear differences ranged from −0.054 to 0.045 percentage points, and the largest absolute difference was for the integrated score's parental-adjusted association with any chronic pain. For atopic dermatitis (19 cases), vitiligo (37 cases) and testicular cancer (60 cases), the probit family variance converged to its boundary of zero.

**Depression diagnosis in ABCD.** ABCD contributed no discovery data. The prespecified ABCD outcome counted a child as a case if the youth or a parent reported present, past or partially remitted major depressive disorder on the computerized Kiddie Schedule for Affective Disorders and Schizophrenia (K-SADS) (Townsend et al., 2020) at baseline or at the two-, four- or six-year follow-up. All other children with a diagnostic interview were controls. The analysis used the 6,323 White children of the population sample with K-SADS data (1,855 cases, 5,127 families) in the paper's reference score units. Logistic regressions adjusted for sex, age at the last interview and PCs 1–10. Per reference-score SD, the log-odds coefficient was 0.377 for the integrated score and 0.317 for the PGC score (19.0% higher, 95% CI 8.9–29.9%). Liability-$R^2$ was 4.9% (3.2–6.5%) and 3.5% (2.3–4.7%), a relative gain of 39.4% (16.7–66.2%, $P = 0.002$). Intervals used the 40 whole-family jackknife groups of the observed-scale $R^2$ comparisons, with Fieller intervals for ratios. The liability conversion used the sample prevalence (29%) because the cohort was not ascertained on depression, and no repeat correction was applied. Because either informant at any of four interviews could establish a case, and the children had not yet passed through the period of risk, this outcome differs from adult lifetime depression, so only the relative gain is comparable with UK Biobank.

Table S2: UK Biobank depression definition (LifetimeMDD)

| **Component** | **Rule (UK Biobank field)** |
|---|---|
| Lifetime route | At least two weeks of depressed mood (20446) or loss of interest (20441), plus at least five of eight symptom domains: depressed mood (20446), loss of interest (20441), weight change (20536), sleep change (20532), tiredness (20449), worthlessness (20450), difficulty concentrating (20435) and thoughts of death (20437). "Yes" counts as present, and weight gain, weight loss or both count as weight change. The questionnaire has no psychomotor item. No additional requirement on the fraction of the day or the frequency of depressed days (20436, 20439) was imposed. |
| Current route | All nine current-symptom items answered, with at least five present "nearly every day", including depressed mood or loss of interest: depressed mood (20510), loss of interest (20514), appetite (20511), sleep (20517), psychomotor change (20518), tiredness (20519), feelings of inadequacy (20507), concentration (20508) and thoughts of suicide or self-harm (20513). This is an item count, not a PHQ sum-score cutoff. |
| Impairment | Both routes also require "a lot" of interference with normal roles during the worst lifetime episode (20440), as in the original definition (Cai et al., 2020). There is no separate measure of current impairment. |
| Case exclusions | Otherwise qualifying cases are left unclassified if they self-reported schizophrenia (20002 code 1289), mania or bipolar disorder (1291), alcohol dependency (1408), opioid dependency (1409) or other substance abuse or dependency (1410), or if field 20126 indicates bipolar I or II disorder. As in the original definition, these exclusions apply only to cases. |
| Controls | An explicit "No" to prolonged depressed mood (20446) and failure to meet the symptom-plus-impairment rule. A negative answer on loss of interest is not required, and the case exclusions are not applied to controls. The original publication describes controls only as participants without the case criteria. This rule matches the size of the published control group to within 2% (Supplementary Note 2). |
| Missing answers | Missing or declined answers never count as symptoms. The lifetime route counts the symptoms observed without requiring every follow-up item, whereas the current route requires all nine items. A missing impairment answer cannot make a case, and a missing mood answer cannot make a control. Participants who meet neither definition, including excluded cases, remain unclassified. |
| Repeat questionnaire | The same rules were applied to the 2022–2023 questionnaire using the matching fields (first-questionnaire field in parentheses): 29011 (20446), 29012 (20441), 29021 (20536), 29022 (20532), 29018 (20449), 29027 (20450), 29026 (20435), 29029 (20437), 29031 (20440), 29003 (20510), 29002 (20514), 29006 (20511), 29004 (20517), 29009 (20518), 29005 (20519), 29007 (20507), 29008 (20508) and 29010 (20513). In the repeat questionnaire, weight-change codes 0, 1 and 2 denote a change (3, no change) and impairment code 0 denotes "a lot". Case exclusions and eligibility were held fixed across the two questionnaires. |

Table S3: UK Biobank depression: liability-scale summaries, score comparisons and stability of the classification

**A. Approximate liability-$R^2$, % (95% CI)**

| | Population | | Implied within-family | |
|---|---|---|---|---|
| Specification | Integrated | PGC | Integrated | PGC |
| Uncorrected | 7.0 (5.5, 8.7) | 4.6 (3.4, 6.1) | 7.4 (4.6, 11.2) | 4.8 (2.7, 7.9) |
| Binary kappa | 8.5 (6.7, 10.5) | 5.6 (4.2, 7.4) | 9.0 (5.6, 13.5) | 5.9 (3.3, 9.6) |
| Polychoric | 9.1 (7.2, 11.3) | 6.1 (4.5, 7.9) | 9.6 (6.0, 14.5) | 6.3 (3.5, 10.3) |
| Stability mixture | 9.3 (7.4, 11.5) | 6.2 (4.6, 8.1) | 9.9 (6.1, 14.8) | 6.5 (3.6, 10.6) |

**B. Paired integrated-minus-PGC comparisons (95% CI)**

| | Population | | Implied within-family | |
|---|---|---|---|---|
| Specification | Difference, pp | Relative gain, % | Difference, pp | Relative gain, % |
| Uncorrected | 2.35 (1.44, 3.35) | 50.8 (28.2, 79.8) | 2.58 (0.73, 4.64) | 53.4 (13.4, 114.8) |
| Binary kappa | 2.82 (1.73, 4.02) | 50.0 (27.8, 78.5) | 3.10 (0.86, 5.58) | 52.6 (12.9, 113.5) |
| Polychoric | 3.01 (1.85, 4.28) | 49.7 (27.6, 78.0) | 3.31 (0.90, 5.96) | 52.3 (12.7, 113.0) |
| Stability mixture | 3.07 (1.89, 4.37) | 49.6 (27.6, 77.8) | 3.38 (0.92, 6.08) | 52.1 (12.6, 112.8) |

**C. Repeat-status transitions**

| | Repeat assessment, 2022–2023 | | | |
|---|---|---|---|---|
| Initial assessment, 2016–2017 | Control | Unclassified | Case | Total |
| Control | 31,052 | 4,253 | 505 | 35,810 |
| Unclassified | 11,605 | 17,484 | 2,606 | 31,695 |
| Case | 1,559 | 4,494 | 5,514 | 11,567 |
| Total | 44,216 | 26,231 | 8,625 | 79,072 |

**D. Repeat-stability inputs**

| Correction | Repeat $N$ | Input $s$ (95% CI) | Calibration |
|---|---|---|---|
| Binary kappa | 38,630 | 0.810 (0.802, 0.818) | Case/control agreement |
| Polychoric | 79,072 | 0.751 (0.747, 0.757) | Single Gaussian copula |
| Stability mixture | 79,072 | 0.733 (0.727, 0.739) | Two-component Gaussian copula |

*Notes.* Panels A and B use the 7,054 participants (1,679 cases) described in Supplementary Note 2, with identical covariates and score units for both scores and an assumed prevalence of 0.15. "Uncorrected" uses $s = 1$, and the other rows divide the log odds ratio by $\sqrt{s}$ before conversion. Within-family values are model-implied from the SNIPar direct-to-population ratio. Panel B gives differences in percentage points and relative gains, $100(R^2_{\text{Integrated}}/R^2_{\text{PGC}} - 1)$. Intervals come from 4,000 paired whole-family bootstrap samples.

Panel C includes all respondents with an interpretable state at both assessments. The median interval between assessments was 6.2 years, and 25,022 respondents (31.6%) changed category. Of the initial cases, 47.7% were cases again, 38.9% were unclassified and 13.5% were controls. Kappa (Panel D) excludes respondents who were unclassified at either assessment, whereas the polychoric and mixture estimates use all three states.

Table S4: Incremental observed-scale variance explained on harmonised samples

| Outcome | $N$ | Added $R^2$, % PGC | Added $R^2$, % Integrated | Gain, % (95% CI) | Outcomes |
|---|---|---|---|---|---|
| UK Biobank: depression | 7,054 | 2.434 | 3.685 | 51.4 (26.7, 90.6) | 1 |
| UK Biobank: internalizing | 9,895 | 2.418 | 4.697 | 94.2 (68.1, 131.1) | 1 |
| ABCD: internalizing | 5,696 | 2.161 | 3.179 | 47.1 (22.5, 81.7) | 1 |
| UK Biobank: anxiety | 11,839 | 1.540 | 2.818 | 82.9 (59.4, 115.1) | 1 |
| UK Biobank: neuroticism | 29,500 | 2.356 | 4.429 | 88.0 (72.6, 106.9) | 1 |
| UK Biobank: 43 adult outcomes | 10,387–36,316 | 0.275 | 0.432 | 57.2 (41.7, 76.5) | 43 |

$N$ is the number of participants (a range for the adult panel). Both scores use identical observations for each outcome. Values are uncorrected observed-scale $R^2$. The adult panel gives equal-weight means across outcomes, and gains compare those means. Intervals are paired 95% Fieller intervals from 40 whole-family deletions. For the 1,854 ABCD phenome outcomes of Table S10, the same jackknife gave mean observed $R^2$ of 0.237% for the PGC score and 0.295% for the integrated score, a 24.7% gain (95% CI 10.8 to 40.5%), and the corresponding Fieller interval from paired family influences (Supplementary Note 5) is 10.8 to 41.4%.

### Supplementary Note 3: Latent internalizing within families

This note concerns the analyses presented in Figure 1B. It describes the symptom items, the factor models, the separation of each score into between-family and within-family components, and the complete-response checks.

**Symptom items.** UK Biobank used 28 items (Table S5): the nine PHQ-9 (Kroenke et al., 2001) and seven GAD-7 (Spitzer et al., 2006) items from the first online mental-health questionnaire (2016–2017), scored 0–3, and the twelve yes/no items of the EPQ-R-S neuroticism scale from the baseline assessment (2006–2010). ABCD used the 30 items of the anxious/depressed, withdrawn/depressed and somatic-complaints scales of the Youth Self-Report (Achenbach and Rescorla, 2001), which the children completed at the 4.5-, 5.5-, 6- and 7-year follow-ups. The YSR was not administered at baseline. Item 91 (suicidal thoughts) was not administered in ABCD. Each ABCD item was averaged over the visits at which the child answered it, and one answer sufficed.

**Factor models.** In the primary hierarchical model, each item loaded on its scale factor (PHQ, GAD and neuroticism in UK Biobank, and the three YSR scales in ABCD), and the three scale factors loaded on a general internalizing factor. In the alternative bifactor model, each item loaded on both the general factor and its scale factor, and all factors were uncorrelated. Neither model allowed correlated residuals. UK Biobank models treated the items as ordinal and were estimated by WLSMV with the theta parameterization, using all available pairs of items. ABCD models treated the averaged items as continuous and were estimated by robust maximum likelihood (MLR) with full-information handling of missing items and clustering by family. The factor structures were chosen pragmatically during exploratory model development, to summarize the variance shared across the symptom items in a single, less noisy outcome, and were then held fixed for all score comparisons. We do not interpret them as the definitive structure of internalizing psychopathology. They serve as a common target for comparing the scores, and the comparisons were essentially unchanged under an alternative bifactor specification (Supplementary Table S6). Fit indices (Table S6C) come from the models without the scores. In ABCD the bifactor model fitted better than the hierarchical model, but the score comparisons were nearly identical under both structures (Table S6A).

**Within-family associations.** Each child's score $Z$ was split into the family mean $Z_B$ and the child's deviation from it, $Z_W = Z - Z_B$. All genetic and covariate paths pointed to the general factor:

$$I_{if} = \beta_W Z_{W,if} + \beta_B Z_{B,f} + \gamma^\top C_{if} + \zeta_{if}.$$

The coefficient $\beta_W$ is the within-family association. Population associations were estimated on the same participants, with the total score in place of its two components. Covariates were sex, birth year and its square (each standardized) and principal components 1–20 in UK Biobank, and sex, mean YSR age and principal components 1–10 in ABCD. Coefficients were divided by the total SD of the latent factor, $\sqrt{\psi + b^\top \Sigma_X b}$, where $\psi$ is the residual variance, $b$ the vector of path coefficients and $\Sigma_X$ the covariance of the predictors. The within-family deviation was not rescaled. Factors were oriented so that higher values mean more symptoms. Family means used every scored sibling in the reference sample (Supplementary Note 1), each weighted equally. An ABCD family could therefore contribute a single child with YSR data whose family mean included the other siblings.

**Confidence intervals.** Due to WLSMV, in UK Biobank, families were divided into 40 groups. Each group was left out in turn and all parameters were re-estimated, with the score, item and covariate transformations held fixed. The covariance matrix was 39/40 times the sum of cross-products of the deviations of the 40 leave-one-group-out estimates, and intervals used the $t$ distribution with 39 degrees of freedom. In ABCD, MLR allowed for intervals using family-level influence functions derived from the likelihood scores. These propagated the uncertainty in the total latent variance, including the empirical covariance of the predictors, with the finite-sample factor $\sqrt{G/(G-1)}$ for $G$ families. Because both scores were analysed on the same families, the same jackknife groups or influences also gave the covariance needed for paired comparisons. Relative gains were computed on the log-ratio scale. The retention comparison (Table S7) contrasts $100\,\beta_W/\beta_P$ between the scores, where $\beta_P$ is the population coefficient in the same family sample. Every reported latent model, including each jackknife refit, converged, was identified, had admissible covariance matrices and had a projected gradient below $10^{-3}$.

**Complete-response checks.** The primary analyses used every answered item, which assumes that missing items are missing at random. We therefore repeated the UK Biobank analyses among participants who answered all items of the relevant scale, or all 28 items. Besides internalizing, these checks used single-factor models of the twelve EPQ-R-S items (neuroticism) and of the seven GAD-7 items (anxiety), estimated in the same way with the same covariates and family decomposition, first with all available items and then among complete responders. Completeness was assessed for each participant, so a family could contribute a single complete responder. The family decomposition and all transformations were unchanged (Table S6B).

Table S5: Symptom items of the latent internalizing models

| **Cohort** | **Scale (items)** | **Items (UK Biobank field or YSR item number)** | **Coding** |
|---|---|---|---|
| UK Biobank | PHQ-9 depression (9) | Depressed mood (20510), loss of interest (20514), sleep (20517), tiredness (20519), appetite (20511), feelings of inadequacy (20507), concentration (20508), slowness or restlessness (20518), thoughts of suicide or self-harm (20513) | 0–3 (not at all to nearly every day), 2016–2017 questionnaire |
| UK Biobank | GAD-7 anxiety (7) | Nervousness or anxiety (20506), inability to stop worrying (20509), worrying too much (20520), trouble relaxing (20515), restlessness (20516), irritability (20505), foreboding (20512) | 0–3, 2016–2017 questionnaire |
| UK Biobank | EPQ-R-S neuroticism (12) | Mood swings (1920), miserableness (1930), irritability (1940), hurt feelings (1950), fed-up feelings (1960), nervous feelings (1970), worrier or anxious feelings (1980), tense or "highly strung" (1990), worrying after embarrassment (2000), suffering from "nerves" (2010), loneliness or isolation (2020), guilty feelings (2030) | Yes/no, baseline assessment 2006–2010 |
| ABCD | YSR anxious/depressed (12) | Items 14, 29, 30, 31, 32, 33, 35, 45, 50, 52, 71 and 112 | 0–2, averaged over follow-ups |
| ABCD | YSR withdrawn/depressed (8) | Items 5, 42, 65, 69, 75, 102, 103 and 111 | 0–2, averaged over follow-ups |
| ABCD | YSR somatic complaints (10) | Items 47, 51, 54 and 56a–56g | 0–2, averaged over follow-ups |

All items were scored so that higher values indicate more symptoms. YSR responses (0 not true, 1 somewhat or sometimes true, 2 very or often true) come from the 4.5-, 5.5-, 6- and 7-year follow-ups.

Table S6: Within-family associations with latent internalizing

**A. Integrated and PGC scores under two factor structures**

| Cohort | Factor model | Integrated $\beta$ (95% CI) | PGC $\beta$ (95% CI) | Gain, % (95% CI) |
|---|---|---|---|---|
| UK Biobank | Hierarchical | 0.272 (0.244, 0.300) | 0.213 (0.183, 0.242) | 27.8 (17.4, 39.2) |
| | Bifactor | 0.264 (0.237, 0.290) | 0.208 (0.180, 0.236) | 26.8 (16.7, 37.8) |
| ABCD | Hierarchical | 0.269 (0.173, 0.366) | 0.175 (0.075, 0.274) | 54.2 (8.5, 119.2) |
| | Bifactor | 0.266 (0.171, 0.361) | 0.172 (0.076, 0.268) | 54.4 (9.4, 118.1) |

**B. UK Biobank: complete-response analyses**

| Outcome | Items required | $N$/families | Integrated $\beta$ (95% CI) | PGC $\beta$ (95% CI) | Gain, % (95% CI) |
|---|---|---|---|---|---|
| Internalizing | All 28 items | 9,895/8,055 | 0.234 (0.192, 0.275) | 0.185 (0.141, 0.230) | 26.2 (9.0, 46.2) |
| Neuroticism | Available items | 36,292/17,656 | 0.217 (0.193, 0.240) | 0.162 (0.140, 0.185) | 33.4 (23.9, 43.6) |
| | All 12 items | 29,500/16,952 | 0.226 (0.200, 0.252) | 0.166 (0.139, 0.193) | 36.5 (25.2, 48.7) |
| | All 28 items | 9,895/8,055 | 0.202 (0.155, 0.249) | 0.142 (0.097, 0.186) | 42.9 (20.2, 69.9) |
| Anxiety | Available items | 12,000/9,340 | 0.186 (0.143, 0.229) | 0.155 (0.109, 0.200) | 20.3 (0.8, 43.5) |
| | All 7 items | 11,839/9,240 | 0.187 (0.143, 0.231) | 0.152 (0.105, 0.199) | 23.3 (2.8, 48.0) |
| | All 28 items | 9,895/8,055 | 0.172 (0.125, 0.218) | 0.144 (0.096, 0.192) | 19.4 (−3.3, 47.4) |

**C. Fit of the measurement models without scores**

| Cohort | Factor model | CFI | TLI | RMSEA | SRMR |
|---|---|---|---|---|---|
| UK Biobank | Hierarchical | 0.952 | 0.948 | 0.038 | 0.063 |
| | Bifactor | 0.945 | 0.936 | 0.043 | 0.064 |
| ABCD | Hierarchical | 0.839 | 0.826 | 0.064 | 0.058 |
| | Bifactor | 0.910 | 0.896 | 0.050 | 0.040 |

*Notes.* Coefficients are standardized within-family associations (Supplementary Note 3), and gains are $100(\beta_{\text{Integrated}}/\beta_{\text{PGC}} - 1)$. Within each cohort, both scores and both factor models use the same participants: in UK Biobank, 36,296 participants from 17,656 families (the available-item sample, whose internalizing row is the hierarchical row of Panel A), and in ABCD, 1,172 children from 593 families. In ABCD, the hierarchical-model difference between the scores was 0.095 (95% CI 0.033 to 0.156, $P = 0.0025$). In Panel B, neuroticism and anxiety are single-factor models of their scale items, and rows other than the available-item rows require complete answers to the items of each scale or to all 28 items, with completeness assessed per participant. CFI, TLI and RMSEA are scaled fit indices.

Table S7: Share of the population association retained within families

| Outcome | Integrated, % | PGC, % | Difference, percentage points (95% CI below) |
|---|---|---|---|
| UK Biobank: depression | 103.2 | 102.3 | 0.9<br>(−13.7, 14.3) |
| ABCD: internalizing | 130.6 | 93.3 | 37.3<br>(4.2, 70.3) |
| UK Biobank: internalizing | 92.1 | 97.4 | −5.2<br>(−12.5, 2.1) |
| UK Biobank: neuroticism | 94.0 | 96.2 | −2.3<br>(−7.6, 3.1) |
| UK Biobank: anxiety | 89.0 | 100.9 | −12.0<br>(−28.2, 4.3) |
| ABCD: phenome profile | 100.8 | 90.2 | 10.6<br>(−10.9, 32.2) |

Retention is $100\,\beta_{\text{within}}/\beta_{\text{population}}$ in the same family sample, and values above 100% indicate a larger within-family association. Differences subtract PGC from integrated retention, with paired 95% confidence intervals. For UK Biobank depression, the per-score 95% intervals were 83.1–123.3% (integrated) and 78.1–126.5% (PGC). Depression uses the SNIPar direct and population coefficients with the 4,000-sample family bootstrap (Supplementary Note 2), latent outcomes use the jackknife (UK Biobank) or family influences (ABCD) of Supplementary Note 3, and the phenome row compares the corrected within-family-on-population profile slopes across 1,854 outcomes (Supplementary Note 5).

### Supplementary Note 4: Adult health, life and disease outcomes

This note relates to Figure 1C and Table 2. It defines the 20 life and health outcomes and the 23 diagnoses and summarizes the results for the PGC score.

**Outcomes.** Table S8 defines the life and health outcomes and Table S9 the diagnoses. Every outcome was oriented so that higher values mean greater burden or disadvantage. A participant was a case if any listed hospital diagnosis, self-report or additional rule applied in any available record. Participants without case evidence but with a related condition listed as a control exclusion were set to missing, and all others were controls. The diagnoses therefore describe recorded lifetime history, with no common follow-up window or record-coverage cutoff. The outcome labelled Alzheimer's disease combines hospital diagnoses of Alzheimer's disease (G30), dementia in Alzheimer's disease (F00), vascular dementia (F01), dementia in other diseases (F02) and unspecified organic mental disorder (F09) with the self-reported item "dementia, Alzheimer's disease or cognitive impairment". It therefore describes dementia broadly rather than clinically diagnosed Alzheimer's disease alone.

**Models and tests.** Each outcome was analysed in the sibling validation sample with the two SNIPar models of Supplementary Note 2: a child-only model for the population association and a model adding the parental-score sum for the direct association. Models adjusted for sex, birth year and its square, and principal components 1–20, except that the sex-restricted cancers adjusted for birth year and principal components only. Scores and outcomes were standardized in each outcome's analysed sample, so coefficients are outcome SDs per score SD, and binary diagnoses were analysed as linear outcomes. Two-sided Wald tests used family-clustered robust (HC0) standard errors. The direct-to-population ratio has paired Fieller intervals, which are unbounded when the population association is not clearly different from zero. Benjamini–Hochberg correction covered all 23 diagnoses together, separately for each score and for the population and direct coefficients.

**Results for the PGC score.** Complete results for both scores are in the Supplementary Data. For every life and health outcome, the population coefficient of the PGC score was smaller than that of the integrated score, for example 0.107 versus 0.131 for health burden. The share retained within families was similar for the two scores: the direct-to-population ratio was 0.85 (PGC) versus 0.86 (integrated) for health burden and 0.88 versus 0.87 for any chronic pain. For education and occupational status the ratio was close to zero with both scores.

Table S8: UK Biobank life and health outcomes

| Outcome | UKB fields | Definition and orientation |
|---|---|---|
| Health burden | 2178 | Poor = 1, fair = 2, good = 3, excellent = 4. Response mean, reversed. |
| Health dissatisfaction | 4548 | Satisfaction coding below. Response mean, reversed. |
| Tiredness | 2080 | Not at all = 1, several days = 2, more than half the days = 3, nearly every day = 4. Response mean. |
| Financial dissatisfaction | 4581 | Satisfaction coding below. Response mean, reversed. |
| Lower happiness | 4526 | Satisfaction coding below. Response mean, reversed. |
| Family dissatisfaction | 4559 | Satisfaction coding below. Response mean, reversed. |
| Friendship dissatisfaction | 4570 | Satisfaction coding below. Response mean, reversed. |
| Work dissatisfaction | 4537 | Satisfaction coding below. Response mean, reversed. |
| Any chronic pain | 2956, 3404, 3414, 3571, 3741, 3773, 3799, 4067, 6159 | 1 if any of the eight chronic-pain indicators is positive, 0 if at least one is observed and none is positive, and missing otherwise. |
| Number of pain sites | Same eight pain fields, 6159 | Number of positive chronic-pain indicators, ignoring missing ones. Missing if none is observed. Includes general (all-over) pain. |
| Neck/shoulder pain | 3404, 6159 | Site-specific chronic-pain rule (see notes). |
| Hip pain | 3414, 6159 | Site-specific chronic-pain rule (see notes). |
| Back pain | 3571, 6159 | Site-specific chronic-pain rule (see notes). |
| Abdominal pain | 3741, 6159 | Site-specific chronic-pain rule (see notes). |
| Knee pain | 3773, 6159 | Site-specific chronic-pain rule (see notes). |
| Headache | 3799, 6159 | Site-specific chronic-pain rule (see notes). |

| Outcome | UKB fields | Definition and orientation |
|---|---|---|
| Lower fluid intelligence | 20016, 20128 | Raw fluid-intelligence score at the earliest available assessment (the first instance when assessments tied). Participants who attempted at most three questions in every instance were excluded. Values were multiplied by $-1$ so that higher values indicate lower ability. |
| Lower education | 6138 | Highest qualification converted to years: university degree 20, NVQ/HND/HNC 19, other professional qualification 15, A or AS levels 13, O levels/GCSE/CSE 10 and none 7. Missing if declined or unrecognized. Values were multiplied by $-1$ so that higher values indicate lower educational attainment. |
| Lower occupational status | 20277 | SOC2000 occupation code converted to the sex-specific CAMSIS occupational-status scale (male scale for men, female scale for women). Values were multiplied by $-1$ so that higher values indicate lower occupational status. |
| Lower household income | 738 | Household income before tax: less than GBP 18,000 = 1, GBP 18,000–30,999 = 2, GBP 31,000–51,999 = 3, GBP 52,000–100,000 = 4 and greater than GBP 100,000 = 5. Declined answers were missing. Values were multiplied by $-1$ so that higher values indicate lower household income. |

Health, satisfaction and tiredness outcomes are means of the recognized responses over all available assessments, and at least one valid response was required. Satisfaction was coded extremely unhappy = 1, very unhappy = 2, moderately unhappy = 3, moderately happy = 4, very happy = 5 and extremely happy = 6. “Reversed” means multiplied by $-1$, so that every outcome is oriented towards greater burden or disadvantage.

The eight chronic-pain indicators cover general (all-over), neck or shoulder, hip, back, abdominal, knee, head and facial pain. Each field was combined over its available assessments: any “yes” to pain lasting three months or longer counted as pain, and other observed answers counted as none. A site not reported among the pain types of the last month (field 6159) was coded as no pain at that site. A reported site with a missing answer to the three-month question remained missing.

Fluid intelligence used the earliest available assessment. CAMSIS is the published sex-specific occupational-status scale for SOC2000 codes.

Table S9: UK Biobank diagnoses: sources, case codes and control exclusions

| Outcome | UKB fields | Case codes | Additional rules and exceptions |
|---|---|---|---|
| *Psychiatric diagnoses* | | | |
| Bipolar disorder | 20002, 41270, 20126, 20544 | ICD-10: F31, F31[0-9]. Self-report: 1291. | Probable bipolar I/II (20126) or professional mania, hypomania, bipolar or manic-depression diagnosis (20544) also defines a case. |
| Schizophrenia | 20002, 41270, 20544 | ICD-10: F20, F20[0-9]. Self-report: 1289. | Professional schizophrenia diagnosis (20544) also defines a case. F20.4 (post-schizophrenic depression) is included, but F32 depression alone is not a case code. |
| Psychosis spectrum (F20–F29) | 41270, 41202, 41204, 40001, 40002 | ICD-10 prefixes: F20–F29. | Any matching diagnosis, main/secondary diagnosis, underlying death cause or contributory death cause after code normalization. No duration or incident-onset restriction. |

| Outcome | UKB fields | Case codes | Additional rules and exceptions |
|---|---|---|---|
| *Cardiometabolic disease* | | | |
| Coronary artery disease | 20002, 41270, 41272, 6150 | ICD-10: I21, I21[0-49], I22, I22[0189], I23, I23[0-68], I241, I252. Self-report: 1075. OPCS-4: K40[1-489], K41[1-489], K45[1-5], K49[1-489], K502, K75[1-489]. | Field 6150 reporting heart attack also defines a case. |
| Gout | 20002, 41270, 20003 | ICD-10: M10, M10[0-49]. Self-report: 1466. | Allopurinol use at any time (20003, code 1140875408) also defines a case. Control exclusions: ICD-10: M11, M11[01289]. |
| Hypertension | 20002, 41270 | ICD-10: I10. Self-report: 1065, 1072. | I11–I13 and I15 do not qualify as case codes. |
| Type 2 diabetes | 20002, 41270 | ICD-10: E11, E11[0-9]. Self-report: 1223. | — |
| Venous thromboembolism | 20002, 41270 | ICD-10: I26, I26[09], I80, I80[0-389], I82, I82[0-389], O871, O882. Self-report: 1093, 1094, 1068. | — |
| *Immune and inflammatory disease* | | | |
| Asthma | 20002, 41270, 6152 | ICD-10: J45, J45[0189], J46. Self-report: 1111. | Doctor-diagnosed asthma (6152) also defines a case. Control exclusions: ICD-10: J40, J41, J41[018], J42, J43, J43[01289], J44, J44[0189], J47, J980. Self-report: 1113, 1114. |
| Atopic dermatitis | 41270 | ICD-10: L20, L20[089]. | — |
| Inflammatory bowel disease | 20002, 41270 | ICD-10: K50, K50[0189], K51, K51[0-589]. Self-report: 1462, 1463, 1461. | — |
| Multiple sclerosis | 20002, 41270 | ICD-10: G35. Self-report: 1261. | — |
| Psoriasis | 20002, 41270 | ICD-10: L40, L40[0-589]. Self-report: 1453. | — |
| Vitiligo | 20002, 41270 | ICD-10: L80. Self-report: 1661. | — |
| *Cancer* | | | |
| Basal cell carcinoma | 20001, 41270 | ICD-10: C44, C44[0-9]. Self-report: 1061. | C44 also includes nonmelanoma skin malignancies beyond histologically confirmed basal-cell carcinoma. |
| Breast cancer (female) | 20001, 41270 | ICD-10: C50, C50[0-689]. Self-report: 1002. | Restricted to women. |
| Colorectal cancer | 20001, 41270 | ICD-10: C18, C18[0-9], C19, C20. Self-report: 1020, 1022, 1023. | — |
| Melanoma | 20001, 41270 | ICD-10: C43, C43[0-9]. Self-report: 1059. | — |
| Prostate cancer (male) | 20001, 41270 | ICD-10: C61. Self-report: 1044. | Restricted to men. |
| Testicular cancer (male) | 20001, 41270 | ICD-10: C62, C62[019]. Self-report: 1045. | Restricted to men. |
| *Other diagnosed disease* | | | |
| Alzheimer's disease | 20002, 41270 | ICD-10: G30 and G30[0189] (Alzheimer's disease), and F00, F01, F02 and F09 with all subcodes (dementia in Alzheimer's disease, vascular dementia, dementia in other diseases, and unspecified organic mental disorder). Self-report: 1263 (dementia, Alzheimer's disease or cognitive impairment). | The outcome covers dementia broadly, including vascular dementia and cognitive impairment, rather than clinically diagnosed Alzheimer's disease alone. |

| Outcome | UKB fields | Case codes | Additional rules and exceptions |
|---|---|---|---|
| Glaucoma | 20002, 41270, 6148, 6119 | ICD-10: H40, H401, H402, H408, H409. Self-report: 1277. | Reported glaucoma (6148) or any nonmissing affected-eye response (6119) also defines a case. Control exclusions: ICD-10: G453, H17, H17[0189], H18, H18[0-9], H192, H31, H31[0-489], H32, H32[08], H33, H33[0-5], H34, H34[01289], H35, H35[0-9], H36, H36[08], H44, H44[0-9], H45, H45[018], H40[3-6]. |
| Osteoporosis | 20002, 41270 | ICD-10: M80, M80[0-589], M81, M81[0-689], M82, M82[018]. Self-report: 1309. | — |

A participant was a case if any listed code or additional rule applied in any available record. Case evidence took precedence over control exclusions: participants without case evidence but with a listed control-exclusion code were set to missing, and all others, including those with no recorded evidence, were controls. Sex restrictions apply to the named outcomes.

ICD-10 codes came from hospital diagnoses (41270), and self-report codes from noncancer illness (20002) or cancer (20001), as listed. Coronary procedures used OPCS-4 (41272). Psychosis additionally used main diagnoses (41202), secondary diagnoses (41204), underlying death causes (40001) and contributory causes (40002). Other fields are named in the outcome-specific rules. Bracketed digits list the included fourth characters of a code. The outcomes describe recorded history, without a common follow-up period or record-coverage cutoff.

### Supplementary Note 5: Childhood phenome and developmental trajectories

This note relates to Figure 2. It describes how the ABCD outcomes were coded and selected, the association models, the profile slope used to compare sets of associations (also used in Supplementary Notes 6 and 7), the comparison with the PGC score and the trajectory analyses.

**Outcome inventory.** Each ABCD item was analysed separately for each reporter (youth, parent or teacher). When a question was asked only after a positive screening question, children with a negative screen were coded as not having the outcome, and children with a positive screen but a missing follow-up answer were left missing. We excluded items that record details asked only of a subgroup, unordered categorical items, administrative fields, duplicates and items whose question routing was uncertain. The resulting inventory of 2,328 outcomes defines the Benjamini–Hochberg family of the phenome-wide analysis (Figure 2A).

**Association models.** Each outcome was regressed on the score using all assessments of each child (linear probability models for binary outcomes), with sex, baseline age, principal components 1–10, study site and assessment as covariates, and family-clustered (CR1) standard errors. An outcome was analysed if at least 200 children contributed, the outcome varied, and, for binary outcomes, at least 20 children were observed in each category. Outcomes with 20–49 children in a category are flagged as sparse. A child whose status changed between assessments could count in both categories. The sibling analyses used non-twin siblings and dizygotic twins from families with at least two children with usable observations, not necessarily from the same assessment. They comprised a population model fitted to these siblings, which required at least 100 families, and a within-family model with family fixed effects, which required at least 20 families in which the children differed in both outcome and score. Both sibling models required at least ten children in each category of a binary outcome. The family fixed effects were centred within the outcome-specific rows of each analysis. Figure 2A highlights outcomes whose within-family association was nominally significant and in the same direction as the population association. Benjamini–Hochberg-adjusted results are in the Supplementary Data.

**Profile slopes.** To summarize how two sets of association coefficients relate across many outcomes, we used a slope through the origin corrected for sampling error in both sets:

$$\widehat{k} = \frac{\sum_j w_j\{\widehat{x}_j\widehat{y}_j - \widehat{\mathrm{Cov}}(\widehat{x}_j, \widehat{y}_j)\}}{\sum_j w_j\{\widehat{x}_j^2 - \widehat{\mathrm{Var}}(\widehat{x}_j)\}}.$$

Here $x_j$ and $y_j$ are the two association coefficients for outcome $j$, not individual observations, and the sampling variances and covariances come from the fitted models. The slope compares:

- the PGC score ($x$) with the integrated score ($y$),
- sibling population coefficients ($x$) with within-family coefficients ($y$),
- girls ($x$) with boys ($y$),
- main score effects ($x$) with score-by-exposure interactions ($y$),
- White children ($x$) with another population group ($y$).

Each outcome received equal weight ($w_j = 1$) in the primary analyses. Weighting each battery equally was a sensitivity analysis. Every comparison used all outcomes that could be estimated in both models, whatever the strength of their score associations. The slope describes the relative size of two association profiles, not prediction accuracy for individuals.

Uncertainty came from 10,000 wild-bootstrap draws that multiplied each family's linearized influence on the slope by a random sign shared by all members of that family. This preserves the dependence between outcomes, models, sexes and scores. The sampling covariances and reference moments were held fixed. The denominator $D = \sum_j w_j\{\widehat{x}_j^2 - \widehat{\mathrm{Var}}(\widehat{x}_j)\}$ had to be positive. For comparisons of scores, of family models and of population groups, a denominator below 1.96 times its standard error marked a weak reference, and intervals were still reported. Sex profiles required $D > 5\ \mathrm{SE}(D)$.

**Comparison with the PGC score.** Both scores were standardized in the same White reference sample and compared on the identical outcome rows of the 1,854 outcomes that could be estimated in the full population, sibling population and within-family models (Table S10). We also compared the incremental variance explained on the observed scale,

$$\Delta R_{sj}^2 = b_{sj}^2 \frac{\mathrm{SS}_{G_{sj}|C}}{\mathrm{TSS}_{Y_j}},$$

where $b_{sj}$ is the coefficient of score $s$ for outcome $j$, $\mathrm{SS}_{G_{sj}|C}$ the sum of squares of the score after adjustment for the covariates, and $\mathrm{TSS}_{Y_j}$ the total sum of squares of the outcome. In family models, the adjustment included the fixed effects, and the total sum of squares was computed before family centring. Relative gains compare the equal-weight means of the increments. Their intervals are Fieller intervals from paired family influences that propagate the coefficients and the score and outcome variances, and the tests of equal means use the 10,000 shared-family draws. Population and within-family gains are based on different samples and do not measure attenuation. The similarity of the two population profiles is also described by the Pearson correlation of the two scores' coefficients across all 2,328 outcomes. Results for all 2,328 outcomes and for battery-equal weights are in the Supplementary Data.

**Developmental trajectories.** Six outcomes were chosen to illustrate developmental patterns (Table S11). Score deciles were defined once, in all scored White children, independently of the outcomes. Logistic models included the continuous score, assessment and their interaction, together with sex, baseline age, principal components 1–10, study site and the deviation of age from the assessment mean, with family-clustered (CR1) standard errors. An outcome was modelled if it was measured at three or more assessments spanning at least two years of mean age. Each retained assessment needed at least ten children in each outcome category and 100 children in each extreme decile, and the retained assessments together needed at least 30 children in each category. Predicted probabilities for the lowest and highest deciles averaged the model predictions over the observed scores within each decile and a common covariate distribution: each child's earliest complete assessment, with the age deviation set to zero. Mean ages label the assessments. Intervals for the probabilities and risk ratios are pointwise and come from the joint covariance of the logit probabilities and log risk ratios.

Table S10: ABCD phenome: integrated and PGC scores compared on identical outcomes

| *Profile slope of integrated-score on PGC-score coefficients* | | | |
|---|---|---|---|
| Model | Outcomes | Slope | 95% CI |
| Population | 1,854 | 1.11 | (1.04, 1.18) |
| Sibling population | 1,854 | 0.98 | (0.83, 1.14) |
| Within family | 1,854 | 1.10 | (0.84, 1.37) |

The analysis uses the 1,854 outcomes estimable in all three models, weighted equally. Profile slopes are corrected for sampling error, with 95% intervals from 10,000 shared-family multiplier draws. The sibling population and within-family models use the same observations. The correlation between the two scores' population coefficients was $r = 0.988$ across all 2,328 outcomes and $r = 0.990$ across the 1,854 outcomes. Correlated sampling error contributes to these values.

Table S11: Developmental trajectories: outcome definitions and estimates by assessment

| **Assessment** | **Mean age** | **Lowest score decile, %** | **Highest score decile, %** | **Risk ratio (95% CI)** |
|---|---|---|---|---|
| *Current major depression.* Youth report of a current K-SADS diagnosis at each assessment. | | | | |
| Baseline | 10.0 | 0.3 (0.1, 0.7) | 1.1 (0.6, 2.1) | 3.79 (1.14, 12.64) |
| Year 2 | 12.1 | 0.1 (0.0, 0.3) | 1.4 (0.8, 2.4) | 12.50 (4.00, 39.01) |
| Year 4 | 14.2 | 1.1 (0.7, 1.7) | 4.6 (3.2, 6.6) | 4.40 (2.23, 8.68) |
| Year 6 | 16.1 | 1.0 (0.6, 1.6) | 5.0 (3.6, 6.9) | 5.07 (2.65, 9.71) |
| *Current social anxiety.* Youth report of a current K-SADS diagnosis at each assessment. | | | | |
| Baseline | 10.0 | 0.1 (0.0, 0.3) | 0.8 (0.4, 1.6) | 8.16 (2.28, 29.27) |
| Year 2 | 12.1 | 0.3 (0.2, 0.6) | 1.4 (0.8, 2.3) | 4.49 (1.65, 12.24) |
| Year 4 | 14.2 | 1.9 (1.4, 2.7) | 8.3 (6.6, 10.4) | 4.26 (2.69, 6.73) |
| Year 6 | 16.1 | 2.4 (1.8, 3.3) | 7.3 (5.8, 9.3) | 3.01 (1.89, 4.79) |
| *Suicidal thoughts.* Youth and parent K-SADS history of active suicidal thoughts, reported at each assessment. Both reports required, positive if either is positive. | | | | |
| Baseline | 10.0 | 3.9 (3.1, 4.9) | 8.7 (7.2, 10.4) | 2.21 (1.56, 3.13) |
| Year 2 | 12.1 | 4.0 (3.2, 4.9) | 13.7 (11.7, 16.0) | 3.41 (2.47, 4.71) |
| Year 4 | 14.2 | 6.7 (5.6, 8.0) | 23.3 (20.5, 26.4) | 3.48 (2.66, 4.54) |
| *Nonsuicidal self-injury.* Youth and parent K-SADS history of self-injury without suicidal intent. Both reports required, positive if either is positive. | | | | |
| Baseline | 10.0 | 5.0 (4.2, 6.1) | 11.6 (9.9, 13.5) | 2.30 (1.72, 3.09) |
| Year 2 | 12.1 | 3.8 (3.0, 4.8) | 11.5 (9.6, 13.7) | 3.00 (2.12, 4.26) |
| Year 4 | 14.2 | 6.9 (5.7, 8.3) | 20.5 (17.9, 23.5) | 2.98 (2.25, 3.94) |
| *Nicotine or cannabis use.* Youth report of lifetime nicotine and cannabis use by each assessment. Both items required, positive if either is positive. | | | | |
| Baseline | 10.0 | 0.3 (0.1, 0.7) | 0.6 (0.3, 1.3) | 2.34 (0.53, 10.30) |
| Year 1 | 11.0 | 0.8 (0.4, 1.4) | 1.4 (0.8, 2.2) | 1.78 (0.68, 4.69) |
| Year 2 | 12.1 | 1.5 (1.0, 2.2) | 2.9 (2.1, 4.0) | 1.95 (1.05, 3.61) |
| Year 3 | 13.0 | 2.3 (1.7, 3.1) | 6.1 (4.8, 7.7) | 2.62 (1.67, 4.11) |
| Year 4 | 14.2 | 6.0 (5.0, 7.3) | 14.1 (12.1, 16.3) | 2.34 (1.73, 3.15) |
| Year 5 | 15.1 | 10.7 (9.2, 12.4) | 26.7 (24.0, 29.7) | 2.50 (2.00, 3.13) |
| Year 6 | 16.1 | 21.8 (19.6, 24.2) | 42.5 (39.5, 45.6) | 1.95 (1.67, 2.27) |
| Year 7 | 17.0 | 25.1 (22.7, 27.7) | 46.9 (43.7, 50.1) | 1.87 (1.61, 2.16) |
| *Psychiatric inpatient treatment.* Parent report of inpatient psychiatric treatment since the previous assessment (follow-ups 1 to 7). A negative screening answer counts as no treatment, and a positive screen with a missing follow-up answer is missing. | | | | |
| Year 1 | 11.0 | 0.1 (0.0, 0.2) | 1.0 (0.5, 1.9) | 15.71 (3.79, 65.13) |
| Year 2 | 12.1 | 0.2 (0.1, 0.5) | 1.2 (0.6, 2.2) | 6.60 (1.74, 25.07) |
| Year 3 | 13.0 | 0.3 (0.1, 0.6) | 1.5 (0.8, 2.7) | 5.73 (1.63, 20.12) |
| Year 4 | 14.2 | 0.4 (0.2, 0.8) | 2.5 (1.6, 4.0) | 5.83 (2.18, 15.58) |

| Assessment | Mean age | Lowest score decile, % | Highest score decile, % | Risk ratio (95% CI) |
|---|---|---|---|---|
| Year 5 | 15.1 | 0.5 (0.3, 0.8) | 4.1 (2.8, 6.1) | 9.20 (4.24, 19.95) |
| Year 6 | 16.1 | 0.4 (0.2, 0.7) | 4.4 (3.1, 6.4) | 11.80 (5.32, 26.17) |
| Year 7 | 17.0 | 0.8 (0.5, 1.3) | 3.8 (2.7, 5.5) | 4.82 (2.35, 9.88) |

Model-standardized probabilities (with 95% confidence intervals) for children in the lowest and highest deciles of the integrated score, and the ratio of the two. Intervals are pointwise. The outcomes are illustrations chosen by the authors, and the probabilities are not incidence estimates.

### Supplementary Note 6: Sex differences and household moderation

This note supports Figure 3 and Table 3. It describes the sex-specific analyses, the symptom-content ratings, the household exposures and the moderation models, and compares the integrated and PGC scores.

**Sex-specific associations.** Separate models for girls and boys used the same score and outcome scaling as the pooled analyses (the common score SD and the pooled outcome SD) and let the covariate effects differ by sex. Family influences were computed from the original families, so brothers and sisters remained linked. The psychiatric inventory comprised 726 outcomes from the Kiddie Schedule for Affective Disorders and Schizophrenia (K-SADS), the Prodromal Psychosis Scale, the General Behavior Inventory–Mania and the 7-Up Mania items. Benjamini–Hochberg correction covered the 726 boy-minus-girl tests. The direction of each contrast follows the coding of the outcome, so a positive contrast does not necessarily mean a larger absolute association in boys.

The boys-to-girls multiplier $k$ is the profile slope of Supplementary Note 5, with girls' coefficients as $x$ and boys' coefficients as $y$. It was computed for the 726 psychiatric outcomes and for all 2,139 outcomes that could be estimated in both sexes. With $D = \sum_j \{x_j^2 - \widehat{\mathrm{Var}}(x_j)\}$, the linearized influence of family $f$ on the slope is

$$U_{kf} = D^{-1} \sum_j \{(y_j - 2\widehat{k}x_j)U_{xfj} + x_j U_{yfj}\},$$

and the resampling draws were $\widehat{k} + \sum_f \xi_f U_{kf}$ with shared-family random signs $\xi_f$. The same draws gave percentile intervals and centred two-sided tests for the difference between the two scores. In a sensitivity analysis, coefficients and influences were rescaled by the ratio of the pooled to the sex-specific outcome SD, keeping the score units and outcomes unchanged.

**Symptom-content ratings.** To test whether sex differences followed how strongly a symptom is associated with one gender, a language model (`gpt-5.6-sol`) rated the cultural typicality of each psychiatric item among US adolescents aged 12–15, from 1 (strongly typical of boys) through 5 (neutral or mixed) to 9 (strongly typical of girls). Each item was rated in a separate request containing only its wording, reporter and response options, without participant data or association results. Ten items with insufficient, nonspecific or erroneous descriptions were excluded, leaving 716. For each sex, the equally weighted outcome coefficients were regressed on the rating centred at 5. The slope is the change in coefficient per rating point, and the difference between the sexes used the paired family influences. The descriptive curves are local linear (LOESS) fits with tricube weights, a span of 0.70, no robustness iterations and pointwise bands with fixed weights. The ratings and outcome SDs were treated as fixed, so the uncertainty of the ratings is not reflected in the intervals. The ratings describe perceived cultural typicality, not the observed prevalence of symptoms or any individual's gender.

**Household exposures.** Exposures were measured at baseline, from reports that agreed within families.

- *Household resources* combined household income, in its ten ordered categories, with the highest education of the caregiver or partner in five levels (below high school, high school or GED, some college or associate degree, bachelor's degree, postgraduate degree). Both components were standardized across eligible families, averaged with equal weight and standardized again. Both components were required.
- *Material hardship* counted seven unmet needs in the previous year: inability to afford food, lost telephone service, incomplete rent or mortgage payment, eviction, utility shutoff, and forgone medical and dental care. All seven items were required.

- *Residential deprivation* was the scaled weighted sum of the Area Deprivation Index for the baseline address (`le_l_adi__addr1_wsum`, Release 7), not its percentile.

A consistent report from one relative could fill another relative's missing income, education or hardship item. A family was excluded from an exposure if its members disagreed on any required component or if data remained missing. The deprivation index required a value for every family member and agreement across the address measures. Each exposure was standardized across eligible families, with one observation per family, before any outcome restriction. The exposures are therefore family-level baseline measures, not verified shared addresses at each later assessment. Higher values indicate more resources, more hardship and more deprivation.

**Moderation models.** Following Keller (2014), each model included the score, the exposure $E$, $E^2$, the score-by-$E$ interaction, the covariates, and the interactions of both the score and the exposure with every covariate. Latent internalizing models used the broader White population (Supplementary Note 1) with MLR estimation, full-information handling of missing items, family clustering and delta-method intervals based on the total latent SD. They included site indicators, centred on the site distribution of the original reference sample, with their interactions with the score and the moderator. The same site assignments were used for both scores and all moderators. Sex was coded 0 (girls) and 1 (boys). Phenome models also included the site-by-exposure and assessment-by-exposure interactions. In household profiles, the profile slope of Supplementary Note 5 relates the interaction coefficients to the main score coefficients, separately for the symptom and full-phenome inventories. For sex, the reported profile estimate is the boys-to-girls multiplier minus one. Holm correction (Holm, 1979) covered the three household exposures, both for the latent interactions tested against zero and for the symptom-profile tests, whereas outcome-by-exposure tests used Benjamini–Hochberg correction. Main score coefficients refer to an exposure of zero and to average covariate values (centred covariates in the latent models and, in the phenome models, the score coefficient averaged over the covariate values of the analysed observations through the score-by-covariate terms). A latent interaction is the change in the score coefficient, in latent SD units, per SD of the exposure, whereas profile moderation is a proportional change across all outcome coefficients.

**Comparison with the PGC score.** Both scores were compared on identical participants and outcomes. The sex profiles were nearly identical. Across the 726 psychiatric outcomes, $k$ was 0.68 (95% CI 0.53 to 0.82) for the integrated score and 0.68 (0.52 to 0.84) for the PGC score (difference −0.01, −0.10 to 0.09, $P = 0.903$). Across all 2,139 outcomes, the corresponding values were 0.71 (0.57 to 0.84) and 0.70 (0.55 to 0.85), with a difference of 0.00 (−0.09 to 0.09, $P = 0.948$). With within-sex standardization, $k$ was 0.79 for both scores across the psychiatric outcomes and 0.77 and 0.76 across all outcomes (all $P \geq 0.866$). For latent internalizing, the score-by-sex interaction (boys minus girls) was −0.099 for the integrated score and −0.084 for the PGC score (difference −0.015, −0.048 to 0.018, $P = 0.362$). The household interactions were −0.007 and −0.016 for resources, 0.007 and 0.011 for hardship, and 0.016 and −0.007 for deprivation. No difference between the scores was significant (all $P \geq 0.114$, and $P \geq 0.458$ after Holm correction across the four latent contrasts for sex and the three exposures), and none of the household interactions differed from zero (Holm-adjusted $P = 1.000$ for the integrated score and $P \geq 0.943$ for the PGC score). The latent comparisons used 5,720 children from 4,644 families for sex, and 5,311 (4,366 families), 5,609 (4,581) and 5,215 (4,308) children for resources, hardship and deprivation.

### Supplementary Note 7: Portability across population groups

This note is about Figure 4. It defines the population groups, the models for the core outcomes and the phenome, the published expectations shown for context, and the observed-score check.

**Population groups.** In UK Biobank we used the genetic groups defined by Privé et al. (2022) from principal components, which that study labels by country: United Kingdom, Nigeria, China and India. We refer to them as EUR, AFR, EAS and SAS. These labels describe genetic similarity, not nationality or birthplace. The EUR reference was the family reserve (United Kingdom group), which was excluded from all discovery analyses (Supplementary Note 1). The AFR, EAS and SAS groups included all eligible participants with scores, with no sibling requirement. In ABCD we used the parent-reported groups of Supplementary Note 1: White (reference), Hispanic and African American.

The reduction in association is $1 - \beta_g/\beta_{\text{ref}}$, where $\beta_g$ is the coefficient in group $g$ and $\beta_{\text{ref}}$ the coefficient of the same score in the reference group. Coefficients are log odds ratios for depression and latent-SD coefficients for internalizing, with the score standardized within each group. Ratios were not constrained, so a reduction can exceed 100% or fall below zero.

**Depression in UK Biobank.** Depression used the case and control rules of Supplementary Note 2 in all four groups, with the case-specific psychiatric exclusions and without the British or Caucasian restriction. Agreement between reported and genetic sex and the absence of a sex-chromosome aneuploidy flag were required. Models adjusted for sex, birth year and its square, and principal components 1–10. Each group's score moments and analysis rows were fixed, and uncertainty came from 40 whole-family deletion groups shared by both scores, with $t_{39}$ intervals. A group was analysed if it had at least 40 participants, including 20 cases and 20 controls, and flagged as small if it had fewer than 200 participants or fewer than 100 cases or controls. The AFR and EAS depression analyses are small by this rule and are shown with open markers in Figure 4A.

**Latent internalizing.** In ABCD, the latent models used the 30 averaged YSR items with MLR estimation, full-information handling of missing items and no correlated residuals, with item and covariate transformations fixed and each group's own score moments. Children needed at least one answered item, complete sex, age and principal components, and genotype and family quality control. A group needed at least 150 children for the model to be fitted and 500 to be included in the main analysis. Intervals used family influences that propagated the fitted parameters, the empirical covariance of the predictors and the joint covariance of the scores and groups, including families that spanned groups.

In UK Biobank, the latent models used the hierarchical model of Supplementary Note 3 without correlated residuals, WLSMV estimation with the theta parameterization, all available pairs of items, and sex, birth year and its square (each standardized) and principal components 1–20 as covariates. Score moments were computed in each group before any item or covariate restriction. Sparse response categories were merged without reference to the scores: adjacent positive PHQ and GAD categories were combined until each category had at least ten observations in every group, keeping the zero category and the binary neuroticism items. The final coding combined categories 2 and 3 for depressed feelings (field 20510), appetite (20511), concentration (20508), restlessness (20516), irritability (20505) and foreboding (20512), and all positive categories for slowness or restlessness of movement (20518) and thoughts of suicide or self-harm (20513), whereas the other PHQ and GAD items kept all four categories. The coding and all transformations were the same for both scores and in every jackknife refit. All 40 whole-family deletions were refitted, including the empirical covariance of the predictors in the total latent variance, and the joint deletion covariance gave $t_{39}$ coefficient intervals and Fieller ratio intervals. The same model and fit criteria applied to every group and both scores. In every group, and more so in the non-EUR groups, most participants had not completed the online questionnaire, so for most of them the latent factor is informed only by the neuroticism items from the baseline assessment (Table S12C gives the share who answered all 28 items).

**Intervals for reductions.** Fieller intervals were obtained by inverting

$$(\widehat{\beta}_g - r\widehat{\beta}_{\text{ref}})^2 \leq q^2\{V_g + r^2 V_{\text{ref}} - 2rC_{g,\text{ref}}\},$$

where $V$ and $C$ are the sampling variances and covariance, and the reduction interval is $[1 - U_r, 1 - L_r]$. The critical value $q$ came from the $t_{39}$ distribution for the UK Biobank analyses and for the observed-score analyses in both cohorts, and from the normal distribution for the ABCD latent analyses. Unbounded or disjoint Fieller sets are reported as such. Similar standardized coefficients do not show that a measure works in the same way in every group.

**Phenome profiles.** Phenome-wide portability compared each group's outcome coefficients with those of White children using the profile slope of Supplementary Note 5, weighting outcomes equally and including every outcome eligible in both groups (at least 200 children and, for binary outcomes, 20 in each category) whatever its significance. The models used the covariates of the phenome-wide analysis, and the coefficients of both groups were expressed in the score and outcome SDs of White children. Figure S1 shows all points underlying Figure 4B, with the same outcomes, slopes and intervals.

**Published expectations.** The expected reductions shown for context are $1 - \sqrt{R^2_{\text{relative}}}$, where $R^2_{\text{relative}}$ is the variance explained in a group divided by that in the European reference. For UK Biobank we used the relative partial $r^2$ that Privé et al. (2022) estimated across 245 traits: 0.180 for AFR, 0.486 for EAS and 0.647 for SAS (expected coefficient retention 0.424, 0.697 and 0.804). For ABCD we used the corresponding estimates of Moore et al. (2025), as applied to ABCD by Wolfram et al. (2026). For the Hispanic group, a linear model of these estimates on genetic distance predicts a relative $R^2$ of 0.740 at the distance of 0.32 that Wolfram et al. (2026) used for admixed American populations, based on Ding et al.

(2023). For African American children we interpolated between the relative $R^2$ of 0.283 that Moore et al. (2025) found in Nigerian-ancestry individuals and 1 for Europeans, $0.8 \times 0.283 + 0.2 \times 1 = 0.4264$, assuming 80% African ancestry. That proportion is an assumption of Wolfram et al. (2026), not a measurement in ABCD. The expected coefficient retention is therefore 0.860 and 0.653. These expectations come from other traits, were not calibrated to depression and carry no uncertainty. We did not test observed against expected values. The relative $R^2$ compares variance explained within each group, which corresponds to within-group standardization of the score and outcome. It therefore matches the core-outcome reductions in Table S12A, but the phenome profile slopes use the score and outcome SDs of White children, so their comparison with the expected slopes assumes similar SDs in the compared groups.

**Observed-score check.** To see whether the latent measurement model drove the portability results, we repeated the core comparisons with observed symptom scores. In ABCD, the outcome was the mean of the raw anxious/depressed, withdrawn/depressed and somatic-complaints sums, averaged over the visits with complete answers. In UK Biobank, it was the mean of the raw PHQ, GAD and neuroticism sums among participants who answered all 28 items. A second version prorated each scale when at least 8 of 9 PHQ, 6 of 7 GAD and 10 of 12 neuroticism items were answered. Score units and covariates were unchanged, and each outcome definition had its own reference-group denominator. Observed coefficients use the empirical SD of the composite, whereas latent coefficients use the model-implied SD. Forty whole-family deletion refits re-estimated the outcome SDs, and pointwise 95% Fieller intervals used $t_{39}$ and required every deletion to be estimable (Table S12C and Figure S2). The ABCD composites covered more than 99% of each group and broadly reproduced the latent pattern. The UK Biobank comparisons are more restricted, because most participants in every group lacked complete PHQ and GAD answers. In the SAS group, the change from the latent to the observed-score reduction came mainly from the reference-group denominator. Because the sample, the outcome and the inference all change between the latent and observed analyses, the difference cannot be attributed to the measurement model alone.

Table S12: Portability of the integrated and PGC scores across population groups

| *A. Reduction of the core association relative to the reference group, % (95% CI)* | | | | |
|---|---|---|---|---|
| Group | Participants | Integrated | PGC | Expected |
| *UK Biobank: depression* | | | | |
| AFR | 374 | 61.9 (8.7, 111.0) | 60.1 (−11.5, 127.0) | 57.6 |
| EAS | 281 | 71.2 (−6.1, 146.8) | 31.5 (−69.2, 132.0) | 30.3 |
| SAS | 581 | 15.7 (−19.6, 50.2) | −19.5 (−69.6, 26.8) | 19.6 |
| *UK Biobank: latent internalizing* | | | | |
| AFR | 4,015 | 57.0 (37.6, 75.9) | 48.9 (16.2, 79.8) | 57.6 |
| EAS | 1,803 | 30.0 (−4.7, 62.6) | 28.8 (−22.2, 76.9) | 30.3 |
| SAS | 6,507 | 17.3 (−1.1, 34.8) | 13.1 (−9.8, 35.1) | 19.6 |
| *ABCD: latent internalizing* | | | | |
| Hispanic | 2,031 | 27.0 (−2.5, 53.8) | 23.2 (−15.1, 57.0) | 14.0 |
| African American | 1,491 | 67.2 (35.1, 98.1) | 89.7 (49.3, 129.5) | 34.7 |
| *B. ABCD phenome: profile slope relative to White children (95% CI)* | | | | |
| Group | Outcomes | Integrated | PGC | Expected |
| Hispanic | 2,147 | 0.78 (0.61, 0.95) | 0.79 (0.60, 0.98) | 0.860 |
| African American | 2,102 | 0.65 (0.38, 0.91) | 0.58 (0.29, 0.85) | 0.653 |
| *C. Observed-score check: standardized coefficients (95% CI) and coverage* | | | | |
| Group | Participants (% of eligible) | Integrated | PGC | |
| *ABCD: mean of the three YSR scale sums* | | | | |
| White | 5,696 (99.6%) | 0.182 (0.154, 0.210) | 0.151 (0.123, 0.180) | |
| Hispanic | 2,015 (99.2%) | 0.123 (0.081, 0.166) | 0.107 (0.055, 0.159) | |
| African American | 1,483 (99.5%) | 0.055 (−0.005, 0.115) | 0.017 (−0.034, 0.068) | |
| *UK Biobank: mean of the PHQ, GAD and neuroticism sums, all 28 items answered* | | | | |
| EUR | 12,028 (26.3%) | 0.213 (0.193, 0.233) | 0.156 (0.137, 0.175) | |
| AFR | 383 (9.5%) | 0.078 (−0.022, 0.179) | 0.055 (−0.027, 0.137) | |
| EAS | 312 (17.3%) | 0.088 (−0.034, 0.209) | 0.101 (−0.040, 0.242) | |
| SAS | 626 (9.6%) | 0.244 (0.170, 0.319) | 0.232 (0.154, 0.310) | |

*Continued on next page*

*Portability (continued)*

| | | | |
|---|---|---|---|
| *UK Biobank: as above, at least 80% of each scale's items answered* | | | |
| EUR | 14,057 (30.7%) | 0.219 (0.201, 0.237) | 0.163 (0.147, 0.178) |
| AFR | 490 (12.2%) | 0.096 (0.011, 0.182) | 0.086 (−0.004, 0.177) |
| EAS | 402 (22.3%) | 0.110 (−0.004, 0.224) | 0.114 (−0.014, 0.242) |
| SAS | 785 (12.1%) | 0.221 (0.143, 0.299) | 0.186 (0.104, 0.267) |

Panel A gives $100(1-\beta_g/\beta_{\text{ref}})$, where each score is standardized within the group and compared with its own reference-group coefficient (UK Biobank EUR, ABCD White children). Depression uses log odds ratios and internalizing standardized latent coefficients. Intervals are pointwise 95% Fieller intervals. The UK Biobank AFR and EAS depression samples include 49 and 54 cases. Panel B uses the score and outcome SDs of White children for both groups, with profile slopes corrected for sampling error and 95% multiplier intervals. "Expected" values are derived from published relative variance explained for other traits (Supplementary Note 7). They carry no uncertainty and are shown for context. Panel C gives observed-score coefficients per group-specific score SD with 95% jackknife intervals. Coverage is relative to each group's sample before any item-completeness requirement. Figure S2 shows the corresponding reductions.

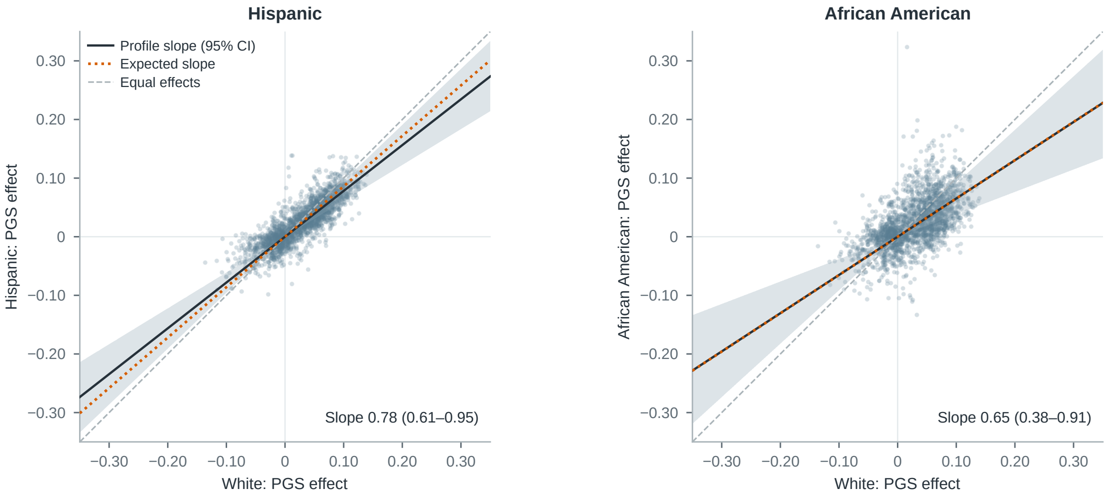


Figure S1: **Full-range phenome portability profiles.** Associations of the integrated score with every eligible ABCD outcome in Hispanic (left) and African American children (right), plotted against the associations in White children. All coefficients are in White-group score and outcome SD units. Figure 4B shows the same outcomes, slopes and intervals on a narrower range. Solid lines show the profile slope with its 95% confidence band, dashed lines equal effects and dotted lines the expected slope (Supplementary Note 7).

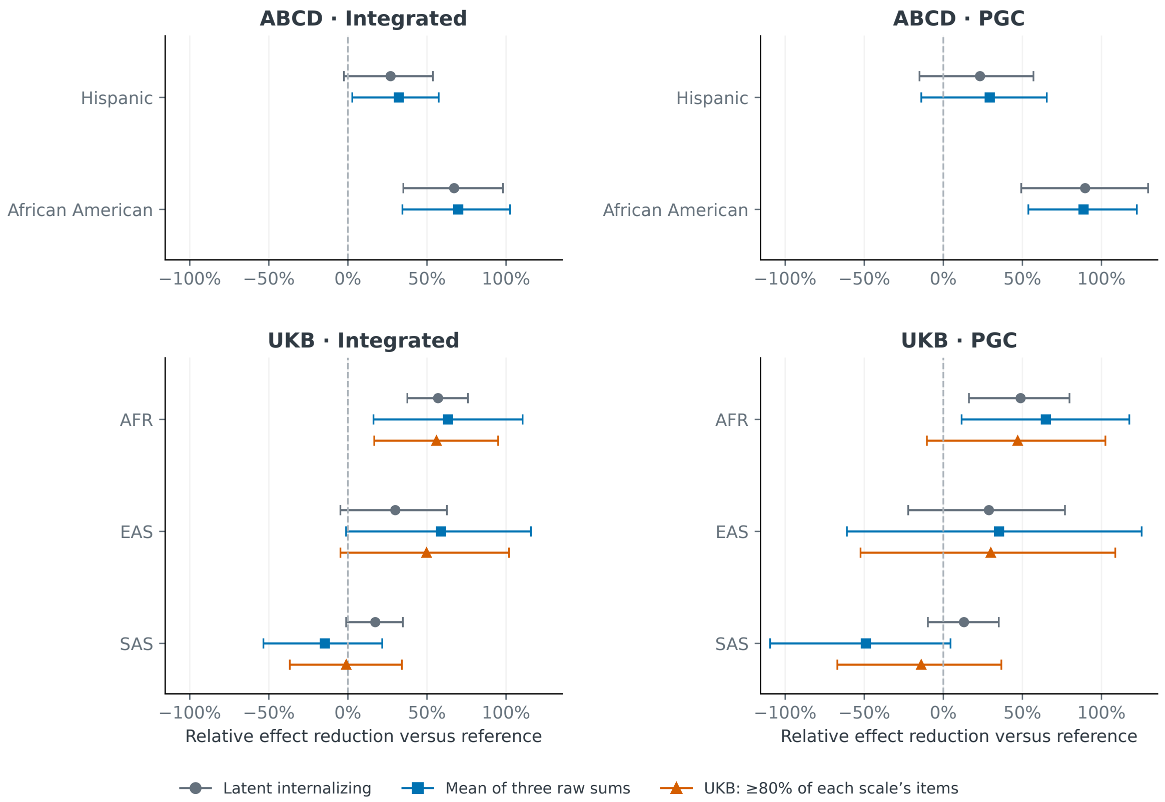


Figure S2: **Observed-score check of portability.** Reduction in association relative to the reference group, $1 - \beta_g/\beta_{\text{ref}}$, for the integrated and PGC scores (reference: White children in ABCD, EUR in UK Biobank). Grey circles show latent internalizing, blue squares the mean of the three observed scale sums, and orange triangles the UK Biobank composite that allows a few unanswered items. Bars are pointwise 95% Fieller intervals. Negative values indicate a larger association than in the reference group. Table S12C gives the coefficients and sample coverage.

### Supplementary References


ABCD Study. ABCD 7.0 data release: Release notes. https://docs.abcdstudy.org/v/7_0_2/documentation/release_notes/7_0.html, 2026. Release 13 May 2026; accessed 21 September 2026.

Thomas M. Achenbach and Leslie A. Rescorla. *Manual for the ASEBA School-Age Forms & Profiles*. University of Vermont, Research Center for Children, Youth, & Families, Burlington, VT, 2001. ISBN 978-0-938565-73-4. URL https://store.aseba.org/MANUAL-FOR-THE-ASEBA-SCHOOL-AGE-FORMS-PROFILES/productinfo/505/.

Mark J. Adams, Fabian Streit, Xiangrui Meng, Swapnil Awasthi, Brett N. Adey, Karmel W. Choi, et al. Trans-ancestry genome-wide study of depression identifies 697 associations implicating cell types and pharmacotherapies. *Cell*, 188(3):640–652.e9, 2025. doi: 10.1016/j.cell.2024.12.002. URL https://doi.org/10.1016/j.cell.2024.12.002.

Brendan Bulik-Sullivan, Hilary K. Finucane, Verneri Anttila, Alexander Gusev, Felix R. Day, Po-Ru Loh, et al. An atlas of genetic correlations across human diseases and traits. *Nature Genetics*, 47(11):1236–1241, 2015. doi: 10.1038/ng.3406.

Na Cai, Joana A. Revez, Mark J. Adams, Till F. M. Andlauer, Gerome Breen, Enda M. Byrne, et al. Minimal phenotyping yields genome-wide association signals of low specificity for major depression. *Nature Genetics*, 52(4):437–447, 2020. doi: 10.1038/s41588-020-0594-5. URL https://doi.org/10.1038/s41588-020-0594-5.

Christopher C Chang, Carson C Chow, Laurent CAM Tellier, Shashaank Vattikuti, Shaun M Purcell, and James J Lee. Second-generation PLINK: rising to the challenge of larger and richer datasets. *Gigascience*, 4(1):7, 2015. doi: 10.1186/s13742-015-0047-8. URL https://doi.org/10.1186/s13742-015-0047-8.

Jacob Cohen. A coefficient of agreement for nominal scales. *Educational and Psychological Measurement*, 20(1):37–46, 1960. doi: 10.1177/001316446002000104. URL https://doi.org/10.1177/001316446002000104.

Andrew Dahl, Michael Thompson, Ulzee An, Morten Krebs, Vivek Appadurai, Richard Border, et al. Phenotype integration improves power and preserves specificity in biobank-based genetic studies of major depressive disorder. *Nature Genetics*, 55(12):2082–2093, 2023. doi: 10.1038/s41588-023-01559-9. URL https://doi.org/10.1038/s41588-023-01559-9.

Yi Ding, Kangcheng Hou, Ziqi Xu, Aditya Pimplaskar, Ella Petter, Kristin Boulier, et al. Polygenic scoring accuracy varies across the genetic ancestry continuum. *Nature*, 618(7966):774–781, 2023. doi: 10.1038/s41586-023-06079-4. URL https://doi.org/10.1038/s41586-023-06079-4.

Sture Holm. A simple sequentially rejective multiple test procedure. *Scandinavian Journal of Statistics*, 6(2):65–70, 1979. URL https://www.jstor.org/stable/4615733.

Longda Jiang, Zhili Zheng, Ting Qi, Kathryn E. Kemper, Naomi R. Wray, Peter M. Visscher, et al. A resource-efficient tool for mixed model association analysis of large-scale data. *Nature Genetics*, 51(12):1749–1755, 2019. doi: 10.1038/s41588-019-0530-8. URL https://doi.org/10.1038/s41588-019-0530-8.

Matthew C. Keller. Gene × Environment Interaction Studies Have Not Properly Controlled for Potential Confounders: The Problem and the (Simple) Solution. *Biological Psychiatry*, 75(1):18–24, 2014. doi: 10.1016/j.biopsych.2013.09.006. URL https://doi.org/10.1016/j.biopsych.2013.09.006.

Kurt Kroenke, Robert L. Spitzer, and Janet B. W. Williams. The PHQ-9: Validity of a brief depression severity measure. *Journal of General Internal Medicine*, 16(9):606–613, 2001. doi: 10.1046/j.1525-1497.2001.016009606.x. URL https://doi.org/10.1046/j.1525-1497.2001.016009606.x.

Sang Hong Lee, Michael E. Goddard, Naomi R. Wray, and Peter M. Visscher. A Better Coefficient of Determination for Genetic Profile Analysis. *Genetic Epidemiology*, 36(3):214–224, 2012. doi: 10.1002/gepi.21614. URL https://doi.org/10.1002/gepi.21614.

Luke R. Lloyd-Jones, Jian Zeng, Julia Sidorenko, Loïc Yengo, Gerhard Moser, Kathryn E. Kemper, et al. Improved polygenic prediction by Bayesian multiple regression on summary statistics. *Nature Communications*, 10(1):5086, 2019. doi: 10.1038/s41467-019-12653-0. URL https://doi.org/10.1038/s41467-019-12653-0.

Ani Manichaikul, Josyf C. Mychaleckyj, Stephen S. Rich, Kathy Daly, Michèle M. Sale, and Wei-Min Chen. Robust relationship inference in genome-wide association studies. *Bioinformatics*, 26(22):2867–2873, 2010. doi: 10.1093/bioinformatics/btq559. URL https://doi.org/10.1093/bioinformatics/btq559.

Spencer Moore, Ivan Davidson, Jonathan Anomaly, Jeremiah H. Li, Mohammad Ahangari, Lauren Moissiy, Michael Christensen, Alexander Strudwick Young, David Stern, and Tobias Wolfram. Development and validation of polygenic scores for within-family prediction of disease risks. *medRxiv*, 2025. doi: 10.1101/2025.08.06.25333145. URL https://www.medrxiv.org/content/10.1101/2025.08.06.25333145v2. Version 2, posted 14 August 2025.

Ulf Olsson. Maximum likelihood estimation of the polychoric correlation coefficient. *Psychometrika*, 44(4):443–460, 1979. doi: 10.1007/BF02296207. URL https://doi.org/10.1007/BF02296207.

Florian Privé, Hugues Aschard, Shai Carmi, Lasse Folkersen, Clive Hoggart, Paul F. O'Reilly, et al. Portability of 245 polygenic scores when derived from the UK Biobank and applied to 9 ancestry groups from the same cohort. *The American Journal of Human Genetics*, 109(1):12–23, 2022. doi: 10.1016/j.ajhg.2021.11.008. URL https://doi.org/10.1016/j.ajhg.2021.11.008.

Robert L. Spitzer, Kurt Kroenke, Janet B. W. Williams, and Bernd Löwe. A brief measure for assessing generalized anxiety disorder: The GAD-7. *Archives of Internal Medicine*, 166(10):1092–1097, 2006. doi: 10.1001/archinte.166.10.1092. URL https://doi.org/10.1001/archinte.166.10.1092.

Lisa Townsend, Kenneth Kobak, Catherine Kearney, Michael Milham, Charissa Andreotti, Jasmine Escalera, et al. Development of Three Web-Based Computerized Versions of the Kiddie Schedule for Affective Disorders and Schizophrenia Child Psychiatric Diagnostic Interview: Preliminary Validity Data. *Journal of the American Academy of Child & Adolescent Psychiatry*, 59(2):309–325, 2020. doi: 10.1016/j.jaac.2019.05.009. URL https://doi.org/10.1016/j.jaac.2019.05.009.

Tobias Wolfram, Spencer Moore, Jeremiah H. Li, Jonathan Anomaly, Ivan Davidson, and Michael Christensen. Interpreting Polygenic Prediction of Cognitive Ability: Evidence for Direct, Reliable, and Portable Genetic Effects. *Intelligence & Cognitive Abilities*, 2(1):1–19, 2026. doi: 10.65550/001c.158459. URL https://doi.org/10.65550/001c.158459.

Alexander I. Young, Seyed Moeen Nehzati, Stefania Benonisdottir, Aysu Okbay, Hariharan Jayashankar, Chanwook Lee, et al. Mendelian imputation of parental genotypes improves estimates of direct genetic effects. *Nature Genetics*, 54(6):897–905, 2022. doi: 10.1038/s41588-022-01085-0. URL https://doi.org/10.1038/s41588-022-01085-0.
